# Unravelling the mechanisms of clozapine-induced neutropenia in treatment-resistant schizophrenia using cellular genomics

**DOI:** 10.64898/2026.11.02.26367124

**Authors:** Yuanhao Yang, Timothy J Edwards, Marie-Odile Parat, Niklas Junker, Sabita Neupane, Panteha Khaledi, Corey Smith, Dan Siskind, Jacob Gratten

**Affiliations:** Mater Research Institute, The University of Queensland, Brisbane, Queensland, Australia; Metro South Addiction and Mental Health Service, Metro South Health, Brisbane, Queensland, Australia; Faculty of Health, Medicine and Behavioural Sciences, The University of Queensland, Brisbane, Queensland, Australia; School of pharmacy and pharmaceutical sciences, The University of Queensland, Brisbane, Queensland, Australia; Translational and Human Immunology Research Laboratory, Infection and Inflammation Program, QIMR Berghofer, Brisbane, Queensland, Australia; Menzies Institute for Medical Research, The University of Tasmania, Hobart, Tasmania, Australia

**Author notes:** These authors contributed equally (Yuanhao Yang, Timothy J Edwards). These authors contributed equally (Dan Siskind, Jacob Gratten).

**Keywords:** schizophrenia, clozapine, clozapine-induced neutropenia, single nucleus RNA sequencing

## Abstract

**Background:** About one-third of individuals with schizophrenia do not respond to first-line antipsychotics. Clozapine has superior efficacy for treatment-resistant schizophrenia (TRS), yet remains underutilised partly due to a 1% risk of potentially fatal clozapine-induced neutropenia (CIN). However, the biological mechanisms underlying CIN remain poorly understood.

**Methods:** We recruited six individuals with TRS who had previously received clozapine: three who ceased clozapine following CIN and three who stopped for reasons unrelated to CIN. Peripheral blood mononuclear cells were collected and cultured *in vitro* under three conditions: untreated, clozapine alone, or clozapine with its two major metabolites. Single nucleus RNA sequencing was performed to profile the cellular transcriptomic landscape following exposure to clozapine and its metabolites.

**Results:** After quality control, 59,313 cells spanning 26 distinct clusters were retained. Multiple cell type-specific genes were differentially expressed in TRS with CIN, or in response to clozapine exposure in TRS donors with CIN (but not in those without CIN), after adjusting for age, sex and smoking status. We also identified a CIN-associated co-expression module in naïve CD4 T cells that was driven by the transcription factor SREBF1, and robust evidence for differential cell-cell communication involving seven intercellular ligand-receptor pairs. There was limited evidence that clozapine-related metabolites contributed to these transcriptional changes.

**Conclusions:** Our findings provide novel insights into cell type-specific transcriptional changes associated with CIN status and/or clozapine exposure in TRS with CIN. With replication in larger, independent cohorts, this work may contribute to the future development of predictive biomarkers for CIN and safer therapeutic strategies for TRS.

## Introduction

Schizophrenia is a chronic and severe mental illness causing persecutory delusions, auditory hallucinations, amotivation and cognitive impairments (1). Symptom onset commonly occurs in early adult life and requires lifelong treatment with antipsychotic medication. Up to one-third of individuals with schizophrenia will develop treatment-resistant schizophrenia (TRS) that is unresponsive to at least two adequate trials of antipsychotic medications (2, 3). Clozapine is the only antipsychotic with established efficacy in TRS, conferring clinically meaningful benefits in approximately 40% of refractory cases (4). Its use is associated with significant improvements in quality of life and reductions in symptom severity, all-cause mortality, suicide risk, and hospitalisation rates (5–9).

Despite its established efficacy, fewer than one in four eligible individuals with TRS are ever prescribed clozapine (10). A major barrier to clozapine initiation is concern regarding its association with serious adverse drug reactions, particularly a rare, unpredictable, and precipitous decline in neutrophil count, leading to severe neutropenia. This haematological adverse effect, termed clozapine-induced neutropenia (CIN), occurs in ∼1% of individuals given clozapine and most cases manifest within 18 weeks of treatment commencement (11, 12). The resultant severe neutropenia confers an increased risk of life-threatening infection, necessitating immediate and often permanent clozapine discontinuation (13). Currently, clozapine is reserved for TRS and requires strict blood monitoring to detect CIN in the first 18 weeks (e.g., weekly) to two years (e.g., 4-weekly) after clozapine initiation (14). Several jurisdictions including New Zealand and those nations covered by the European Medicines Agency have removed mandates for 4-weekly monitoring after the first two years. The United States has historically followed a different monitoring schedule, involving weekly monitoring for the first 6 months, fortnightly monitoring from 6 to 12 months, and monthly monitoring thereafter. Although the FDA formally removed the mandatory clozapine risk evaluation and mitigation strategy in 2025, it continues to recommend absolute neutrophil count monitoring according to the prescribing information (15). Unfortunately, despite intensive surveillance, the rapid onset of CIN after clozapine commencement in some cases may preclude timely detection, highlighting limitations of current monitoring strategies in fully mitigating this haematological risk (16).

Prior genetic studies support a polygenic architecture for CIN, with association signals implicating loci within the human leukocyte antigen (HLA) region and several T cell-related loci (17–20). These findings are consistent with an immune-mediated genetic contribution to CIN susceptibility (21). Pharmacological studies suggest that metabolic bioactivation of clozapine yields reactive intermediates capable of directly injuring neutrophils (22–24). Despite these advances, the biological mechanisms driving CIN remain incompletely defined. A major limitation is that most genomic and mechanistic investigations have relied on bulk□level samples, in which modest cell type-specific genetic or transcriptional signals may be diluted or missed when heterogeneous populations from different hematopoietic hierarchy and immune compartments are analysed in aggregate. To our knowledge, no studies have systematically interrogated the cellular mechanisms underlying CIN and the role of clozapine-related metabolites at single□cell resolution.

Peripheral blood mononuclear cells (PBMCs) provide an accessible surrogate for investigating clozapine-associated immune reactivity and have been used in pharmacogenomic and immunological studies of drug response (25). In this study, we therefore used single nucleus RNA sequencing (snRNA-seq) to profile gene expression in ∼60K high-quality cultured PBMCs obtained from six individuals with TRS (three with CIN and three without), exposed *in vitro* for up to three days to one of three treatments: clozapine alone, clozapine plus its two major metabolites (i.e., N-desmethylclozapine [norclozapine], and clozapine-N-oxide) or untreated. Our goal was to characterise cell type-specific transcriptional signatures associated with CIN status and with response to both clozapine and the combination of clozapine and its metabolites in TRS donors with and without CIN. We identified cell type-specific differentially expressed genes, predominantly in naïve CD4 and CD8 T cells, associated with CIN status and/or exposure to clozapine (and its metabolites) in TRS with CIN, but not TRS without CIN. This specificity extended to broader transcriptional programs, including gene co-expression modules and intercellular communication networks.

## Methods and Materials

### Participant recruitment and blood collection

Participants were recruited from Metro South Addiction and Mental Health Services (MSAMHS), a public mental health service in Queensland, Australia. Eligible participants were recruited if they had a diagnosis of schizophrenia and had a history of previous clozapine treatment. All participants provided written informed consent, and study procedures were conducted in accordance with the National Health and Medical Research Council (NHMRC) National Statement on Ethical Conduct in Human Research (2007). Ethical approval was obtained from Metro South HREC (HREC/2024/QMS/105218). Participants were classified into two groups according to their history of clozapine treatment. The CIN group comprised participants who had experienced an absolute neutrophil count <1,500 cells/µL (<1.50×10^9^/L) at any time during clozapine treatment, with no alternative cause for neutropenia identified. The clozapine-exposed control group comprised participants who had previously received clozapine but had discontinued treatment for reasons other than neutropenia. Individuals were excluded if they were receiving clozapine at the time of recruitment, had a known active haematological malignancy, or had received cytotoxic or myelosuppressive therapy or other significant cytotoxic exposure within the 12 months preceding study screening. For each participant, 20 mL of peripheral venous blood was collected into Acid Citrate Dextrose Solution A (ACD-A)-containing blood collection tubes for subsequent isolation of PBMCs.

### Cell culture

PBMCs were isolated by Ficoll density-gradient centrifugation using Ficoll-Paque PLUS (Cytiva; cat. no. GE17-1440-02). Whole blood was diluted 1:1 (v/v) with phosphate-buffered saline (PBS), layered over 15 mL Ficoll-Paque PLUS, and centrifuged at 400 × g for 40 min at room temperature. Collected PBMCs were resuspended in 50 mL PBS and centrifuged at 400 × g for 10 min at 4 °C. Cells were washed in RPMI 1640 (Gibco, Thermo Fisher Scientific; cat. no. 11875-093), centrifuged again under the same conditions, and resuspended in RPMI 1640.

PBMCs were cryopreserved in 50% RPMI 1640, 40% fetal bovine serum (FBS), and 10% dimethyl sulfoxide (DMSO) using a Mr. Frosty controlled-rate freezing container (Thermo Scientific Nalgene) at −80 °C. Prior to culture, cells were thawed for 1-2 min in a 37 °C water bath, resuspended in RPMI 1640, centrifuged at 400 × g for 10 min at room temperature, and resuspended in complete culture medium consisting of RPMI 1640 supplemented with 10% FBS, 100 U/ml penicillin and 100 μg/ml streptomycin.

PBMCs from each participant were seeded at 1.25×10^6^ cells/mL in 2 mL of culture medium and incubated under one of three conditions: vehicle control (0.003% DMSO), clozapine (1 µM), or clozapine (1 µM) with N-desmethylclozapine (0.5 µM) and clozapine N-oxide (0.2 µM). Clozapine (C6305), N-desmethylclozapine (D5676), and clozapine N-oxide (C0832) were obtained from Sigma-Aldrich. Concentrations were selected based on preliminary cell-viability assays and clinically relevant concentrations of clozapine and its major metabolites (26, 27). Cells were incubated for 68h at 37 °C and 5% CO□, then harvested and washed in culture medium before resuspension for subsequent processing and single nucleus RNA sequencing.

### Single nucleus RNA-seq library preparation and sequencing

Cultured PBMC samples were centrifuged at 350 × g for 5 minutes at room temperature, after which the supernatant was removed, and the pellets were resuspended in 1 mL PBS containing 0.04% BSA (A1595, Millipore Sigma). Cell concentration and viability were assessed using an 8□channel counting slide (ATA Scientific, L72001) and AO/PI reagents (ATA Scientific, F23011) on a Luna□FX7 automated cell counter (Logos Biosystems, L70001).

Cells were fixed for 19 hours at 4□°C following the Fixation of Cells & Nuclei for Chromium Fixed RNA Profiling protocol (10X Genomics, CG000478, Rev C), followed by quenching. Of 18 samples in the study (6 donors × 3 treatment groups), 16 samples were randomly selected for probe hybridization, as the Chromium Fixed RNA Profiling Reagent Kits supports a maximum of 16-plex reactions (10X Genomics, CG000527, Rev F). Hybridization was carried out for 20 hours at room temperature. Post□hybridization nuclei concentrations were assessed using the same AO/PI□based Luna□FX7 workflow.

A single pool of post-hybridisation nuclei was generated by determining the sample with the lowest cell count and combining an equal number of cells from all other samples into a single 15 mL tube. Washing and filtration steps were performed according to the 10X Genomics Protocol CG000527 (Rev F). We then adjusted the final concentration of the library with post-hybridization resuspension buffer to 6,440 nuclei/µL. The final suspension was subsequently loaded onto the 10X Genomics Chromium instrument targeting 128,000 nuclei across the pool of 16 samples, and cell capture and library preparation were performed following the Chromium Fixed RNA Profiling Reagent Kits User Guide. Sequencing was performed at the Australian Genome Research Facility (AGRF) on an Illumina NovaSeq instrument across four S4 flowcell lanes, aiming for a total of 4B reads and >30K reads per nucleus.

### Single nucleus RNA-seq data processing, clustering and annotation

The raw FASTQ sequencing data were initially processed using the Cell Ranger (v9.0.0) “multi” algorithm (28). This pipeline was utilised to demultiplex the sequencing reads per sample, align each sample to the human transcriptome reference, and generate the unique molecular identifier (UMI) count matrix per gene per cell for downstream analyses.

We applied a rigorous data quality control (QC) procedure to each of the 16 individual samples. Initially, cells expressing fewer than 400 genes and genes that were detectable in fewer than 100 cells were excluded. Following this, total cell counts were assessed, and cells were removed if they fell outside the range defined by their median cell count ± 3 times the median absolute deviation (MAD). This criterion was similarly applied to genes, leading to the exclusion of genes with expression levels outside the median ± 3 times the MAD. To eliminate damaged or apoptotic cells, those with a mitochondrial gene percentage (MT%) greater than 5% cells were excluded. The Scrublet (v0.2.3) (29) software was employed to estimate the probability that each cell represented a doublet. We defined the prior probability of the proportion of doublets per sample as 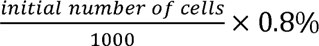 per sample, following the 10X Genomics protocol guidelines for 16-plex runs. We then visually inspected the bimodal distribution of doublet scores per sample and manually adjusted the doublet threshold as needed to differentiate single cells from suspected doublets.

After the implementation of stringent multi-QC steps, the raw UMI count matrices were normalised using the SCTransform (v0.4.1) method (30) for each individual dataset, adjusting for MT% and cell cycle scores as covariates. The normalised data matrices were then integrated utilising the Seurat (v5.2.0) integration workflow (31). Next, unsupervised cell type clustering analysis were performed using the standard Seurat pipeline. Cell types were annotated with the SingleR (v2.2.0) (32) software using recently published scRNA-seq data on 53,981 cultured PBMCs from seven healthy adults as the reference (33). Marker genes were selected based on significant differential expression within the target cluster and were subsequently refined by manual cross□reference to the CellMarker (v2.0) database (34).

### Differential gene expression analysis

Differential gene expression (DE) analysis was conducted (1) between CIN status groups (i.e., “TRS with CIN vs. TRS without CIN”), and (2) within each of the CIN groups across three treatment comparisons: “untreated vs. clozapine”, “untreated vs. clozapine + metabolites”, and “clozapine vs. clozapine + metabolites”. Together, this yielded in a total of seven comparisons (one between group and three within each CIN status group). For each comparison, DE analyses were performed at the individual cluster level. Clusters with fewer than 1% of the total cell number were excluded to maintain sufficient statistical power. The primary DE analysis utilised negative binomial regression, adjusting for covariates including age, sex, smoking status, MT%, and the number of UMIs per cell. For the comparison between CIN status groups, the model also included treatment group and its interaction with CIN status. Significant cluster-specific genes were identified based on the criteria that they were expressed in at least 1% of either group of cells, had an absolute log_2_ fold change (log_2_FC) greater than 0.25, and possessed a Bonferroni-corrected p-value <0.05.

Given the limited number of biological samples recruited in our cohort and the associated risk of false-positive results, two distinct sensitivity analyses were employed to assess the robustness of the primary findings: (i) a non-parametric Wilcoxon rank-sum test, which minimises reliance on the distributional assumptions required by negative binomial regression; and (ii) a generalised negative binomial linear mixed model (implemented using lme4 [v1.1.37] (35)) adjusting for the same covariates described above (fixed effects) and fitting individual as a random effect, to account for non-independence of subject-specific gene expression measures. Significant and high-confidence cluster-specific DE genes were finally determined if they satisfied the Bonferroni-corrected significance thresholds in both sensitivity analyses.

### Co-expression network analysis

High-dimensional weighted gene co-expression network analysis (hdWGCNA [v0.4.9]) (36) was utilised to construct cluster-specific gene co-expression modules in each cluster in which every sample contributed at least 75 cells. Metacells were generated with k□=□25 (number of nearest□neighbour cells pooled to form each metacell) and max_shared□=□10 (maximum number of constituent cells any two metacells may share, limiting overlap and redundancy). Clusters were retained for network inference only if they contained at least 100 metacells per treatment group to improve the stability of gene-gene correlation estimates. The soft□thresholding power was selected as the smallest value at which the scale□free topology model fit plateaued and median connectivity stabilised, which was used to transform the signed gene-gene correlations into a scale□free adjacency matrix. This adjacency matrix was then converted to the topological overlap matrix and subjected to hierarchical clustering to define co□expression modules. For each detected module, the module eigengene was computed as the first principal component of the module’s gene□expression matrix across metacells that served as the module’s representative profile.

To identify cluster-specific gene co-expression modules associated with CIN status and treatment for TRS with CIN and TRS without CIN separately, differential module eigengene (DME) analyses were performed using generalised linear regression, applying the same comparisons and covariate adjustments as in the DE analysis described above. Module eigengenes were normalised using an inverse rank□based transformation. Similarly, two sensitivity analyses were conducted to assess robustness: (i) a non-parametric Wilcoxon rank-sum test and (ii) a generalised linear mixed model. Cluster□specific co□expression modules were deemed significant and high□confidence if they met the Bonferroni□corrected significance threshold in both sensitivity analyses.

For each significant co-expression module identified, transcription factor (TF) regulatory network analysis was performed to elucidate the regulatory architecture underlying the corresponding module. Candidate cis-regulatory elements were mapped by scanning the human genome (hg38) for motifs curated from the JASPAR 2024 CORE vertebrate collection (37). Regulatory networks were inferred using gradient-boosted regression (via XGBoost), in which target gene expression was modelled as a function of TF activity. TF-gene pairs were quantified using regulatory scores derived from the model’s feature importance metrics, reflecting the relative contribution of each TF to predicting its target gene expression while accommodating combinatorial regulation by multiple TFs. Functional regulons were defined at the TF level as sets of high□confidence target genes, retaining TF-gene pairs with regulatory scores greater than 0.05. For each regulon, aggregated activity scores were computed across cells using the RegulonScores algorithm, restricted to target genes showing absolute correlation >0.10 with the corresponding TF. The regulon activity scores were further stratified into positively and negatively co□expressed sub-networks to distinguish transcriptional activation and repression. Activity regulon scores were normalised using an inverse rank□based transformation. Associations between transformed activity regulon scores and CIN/treatment status were evaluated using the same analytical framework (i.e., combining generalised linear regression with sensitivity analyses) as was applied in the DME framework. Module-specific regulons were considered significant if they met the Bonferroni-corrected significance threshold consistently across all models.

### Pathway enrichment analysis

To explore the biological relevance of each significant hdWGCNA module and its associated TFs (with significant regulons), pathway enrichment analysis was performed using enrichR (v3.4) (38). Analyses focused on Gene Ontology (GO) terms, including biological processes (BP), cellular components (CC) and molecular function (MF), with gene set sizes between 15 and 500. Pathways were deemed significant if they had _≥_5 overlapping genes and passed Bonferroni correction. The significant pathways identified by enrichR were further assessed using the Single Cell Pathway Analysis (SCPA [v1.5.2]) method (39) as a sensitivity analysis. Unlike enrichR, which relies on a list of individual genes, SCPA evaluates differences in the multivariate distribution of expression values across all genes in a pathway between conditions. This helps to limit false positive associations from pathways whose signal may be driven by a few genes that are less significant. Pathways were retained as significant if they also had a Bonferroni-corrected SCPA p-value <0.05.

### Cell-cell communication analysis

To assess differences in intercellular communication networks in relation to CIN status and treatment groups, cell-cell communication analysis was performed using the CellChat (v2.2.0) framework (40). Comparisons were made between the CIN groups and separately within each CIN group across three treatment comparisons. This analysis targeted the identification of specific ligand-receptor (L-R) pairs mediating the interactions between cell type clusters that showed significant divergence between treatment groups. Within each cell type cluster, CellChat-based DE genes with p-value <0.05, logFC >0.20, and expressed in >1% cells in at least one group, were used to identify dominant signalling ligands and receptors. For the between CIN status comparison (i.e., “TRS with vs. without CIN”), DE genes were obtained directly from the corresponding DE analysis. For the within group comparison (e.g., untreated vs. clozapine, performed separately within TRS with CIN and TRS without CIN), DE genes were identified in the TRS with CIN, and the same gene set was applied to the TRS without CIN to ensure consistency. For each L-R pair, interaction strength was quantified using the “trimean” method, which robustly estimates the communication probability while minimising the impact of outliers. Analysis was performed across all cell type clusters.

For the comparison between CIN groups, significant intercellular L-R pairs were reported only for cell type clusters containing significant DE genes from the prior DE analysis, and were defined as those with a communication probability >0.01 with permutation p-value <0.001 (after 1,000 permutation tests). For the within group comparisons, significant intercellular L-R pairs were considered specific to TRS with CIN if they (i) were significant in TRS with CIN using the same criteria; and (ii) had a non-significant result in TRS without CIN. Significant L-R pairs specific to TRS without CIN were defined with group assignments reversed.

## Results

### Overall data quality and characteristics

Six participants (aged 22-60 years) with a diagnosis of schizophrenia and a history of previous clozapine treatment were recruited: three were classified into the CIN group (TRS with CIN: 1 female, 2 males), and three into the clozapine-exposed control group (TRS without CIN: 3 males). Participants in the clozapine-exposed control group had ceased clozapine due to adverse effects (sedation, n = 2) and difficulty maintaining long-term adherence (n = 1). Smoking status were also recorded: three participants were smokers (2 TRS with CIN, 1 TRS without CIN) and three were non-smokers (1 TRS with CIN, 2 TRS without CIN). PBMCs were collected from each participant and treated under three conditions: untreated, clozapine, and clozapine + metabolites, yielding a total of 18 donor-treatment combinations (hereafter “samples”). Two samples (i.e., one TRS with CIN and one TRS without CIN, both treated with clozapine) were randomly excluded from the downstream snRNA-seq data generation.

Following the application of stringent QC filters (Figures S1-4), a total of 59,313 single nuclei remained for analysis. Quality assessment of the 16 samples revealed high alignment efficiency (with mapping rates exceeding 96%) and robust feature detection rates (∼10,000 genes per dataset). The post-QC yield of nuclei per sample ranged from 1,800 to 5,600 (Figure S5). Unsupervised clustering and annotation identified 26 distinct cell type clusters, predominantly composed of CD4 T cells, CD8 T cells and natural killer (NK) cells (Figures 1A, 1B and S6-11). Of those, 24 cell type clusters comprised >1% of all nuclei and were retained in downstream analyses, with the exception of CD8 T cell (early memory) and memory CD4 T cell (Th2-like). We observed substantial inter-individual heterogeneity in cellular composition (Figure 1C) and therefore included individual random effects in subsequent analytical models to account for biological variability.

**Figure 1.**
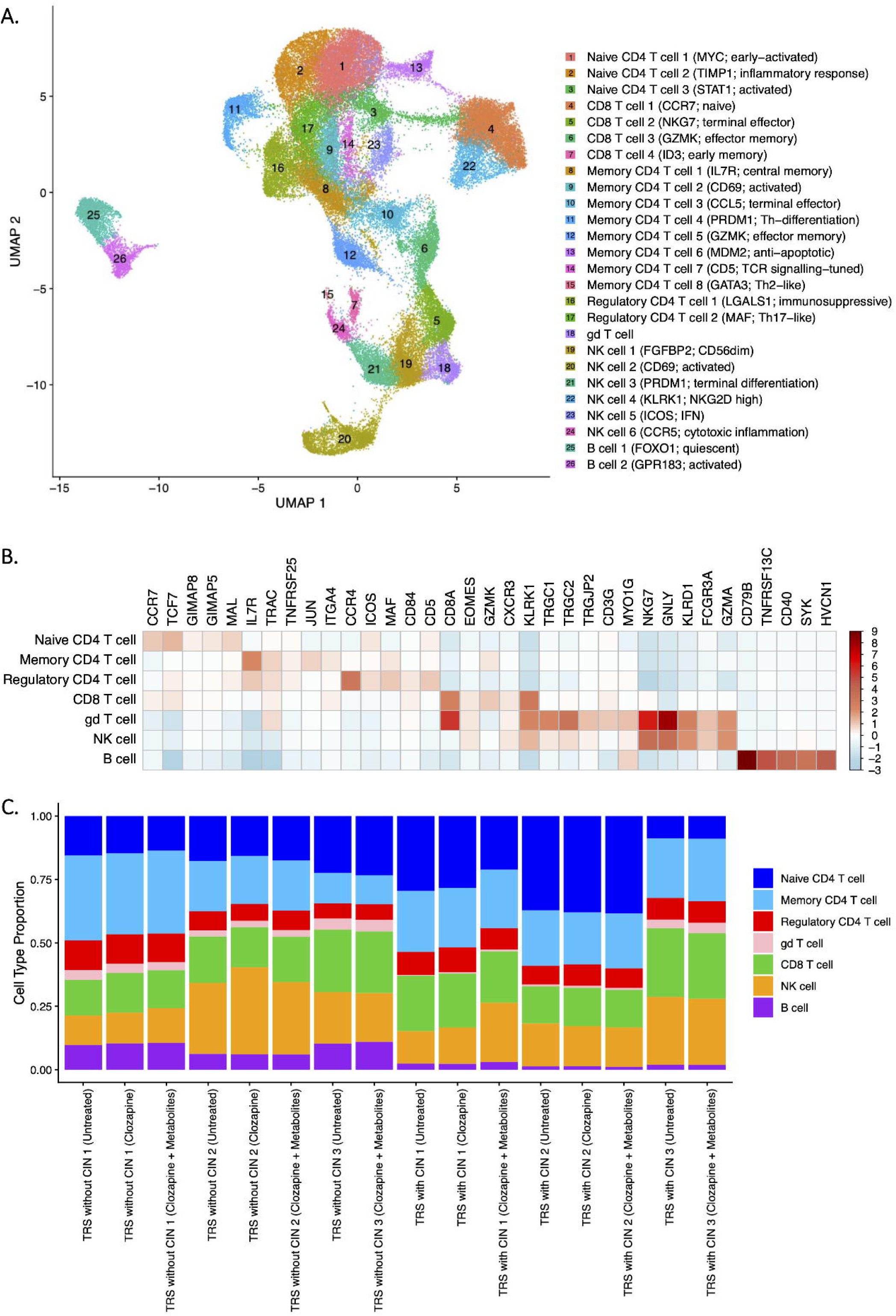
(A) UMAP visualisation of all cells across the 26 clusters. Each cluster is labelled with its cell type annotation and top up-regulated marker genes. (B) Heatmap showing expression of the top five marker genes for each broad cell type. (C) Per-sample distribution of broad cell-type proportions.

### Cell type-specific transcriptional signatures associated with CIN in TRS and response to clozapine and its metabolites

Twenty-four cell clusters met the criteria for cell type-specific differential expression (DE) analysis. Comparing TRS with CIN to TRS without CIN, we identified 40 Bonferroni-significant DE genes across 14 cell type clusters (Figures 2A-B and Table S1), with the majority mapping to naïve CD4 T cells (early-activated; n = 8), CD8 T cells (naïve; n = 7) and NK cells (terminal differentiation; n = 10). Of these 40 associations, 18 genes were found exclusively in one cell type cluster, whereas six genes (*ADTRP*, *EFCAB13*, *FAM118A*, *IL32*, *NDUFA12*, *USP53*) were significant in ≥2 clusters. Notably, *FAM118A* was differentially expressed across multiple CD4 and CD8 T cell types and in one NK cell type (NKG2D-high), with upregulation in TRS donors with CIN (Figure 2C).

**Figure 2.**
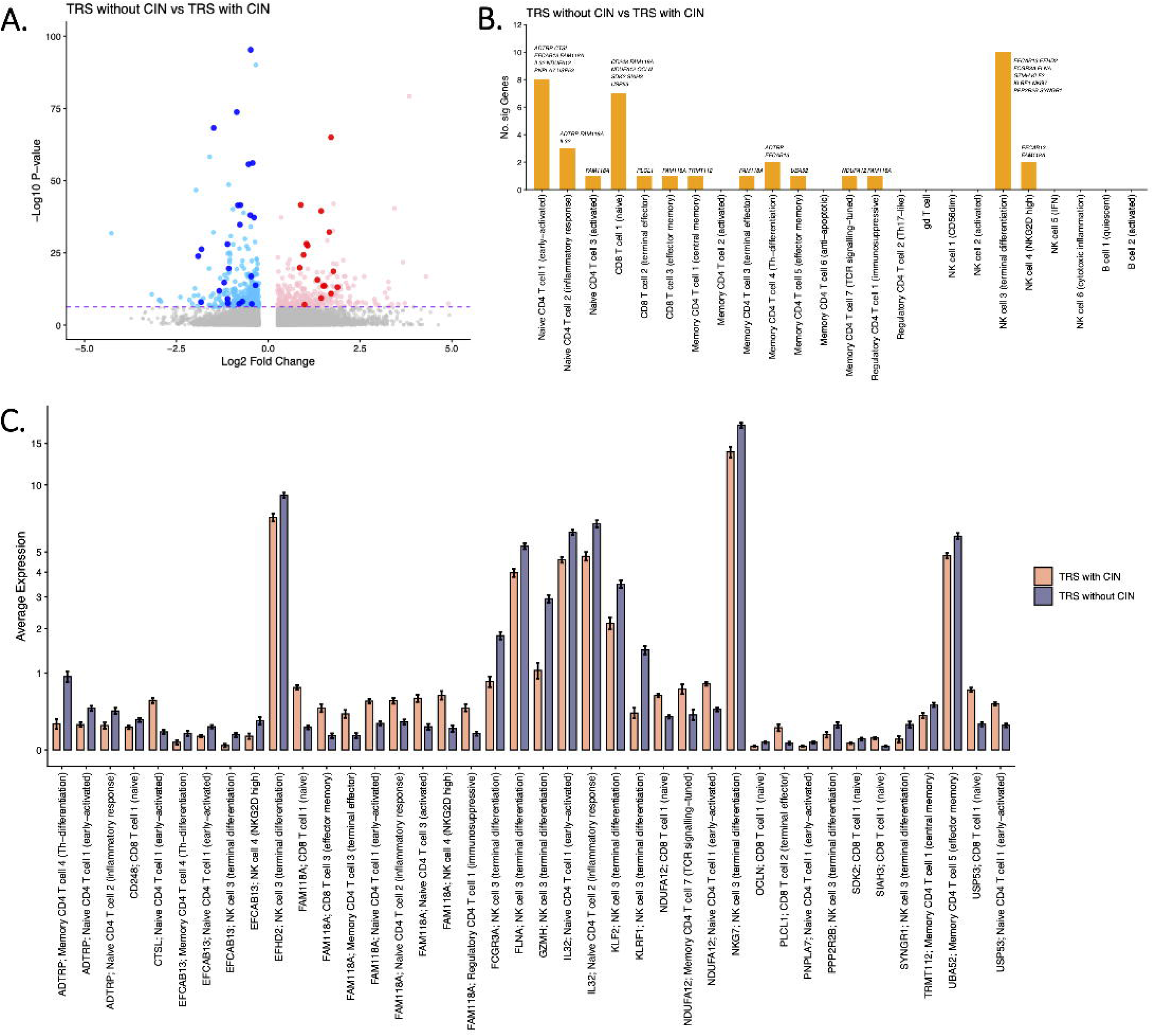
(A) Volcano plot showing differential expression results from analyses comparing TRS with vs without CIN. Significant differentially expressed (DE) genes that passed Bonferroni correction and remained significant in both sensitivity analyses are highlighted. (B) Significant DE genes from panel A, summarised by cluster (number of genes and gene identifiers). (C) Bar plot of average expression for significant DE genes (panel A) in TRS with vs without CIN.

Within each CIN status group, our DE analyses revealed distinct cell type-specific gene expression profiles in TRS with and without CIN (Figures 3A-C and Tables S2-3). In TRS with CIN, clozapine triggered a robust transcriptional response, with 45 genes significantly upregulated relative to untreated samples in specific cell types. These genes were predominantly from naïve CD4 and CD8 T cells, with five genes (i.e., *DDX5*, *EIF1*, *EEF1B2*, *KPNB1*, *UBA52*) overlapping between naïve CD4 T cell (early-activated) and CD8 T cell (naïve), and another three (i.e., *DDX5*, *MT-CO2*, *UBA52*) overlapping among three naïve CD4 T cell types (Figure 3B). In contrast, the transcriptional response to clozapine in TRS without CIN was minimal, with only one gene (*PIK3IP1*) identified as differentially expressed. This gene did not overlap with the cell type-specific expression signatures identified for TRS with CIN. Furthermore, cross-group concordance was weak and non□significant (*ρ* = 0.04, p-value = 0.78) when comparing effect size estimates (log_2_FC) of these significant DE genes between TRS with and without CIN.

**Figure 3.**
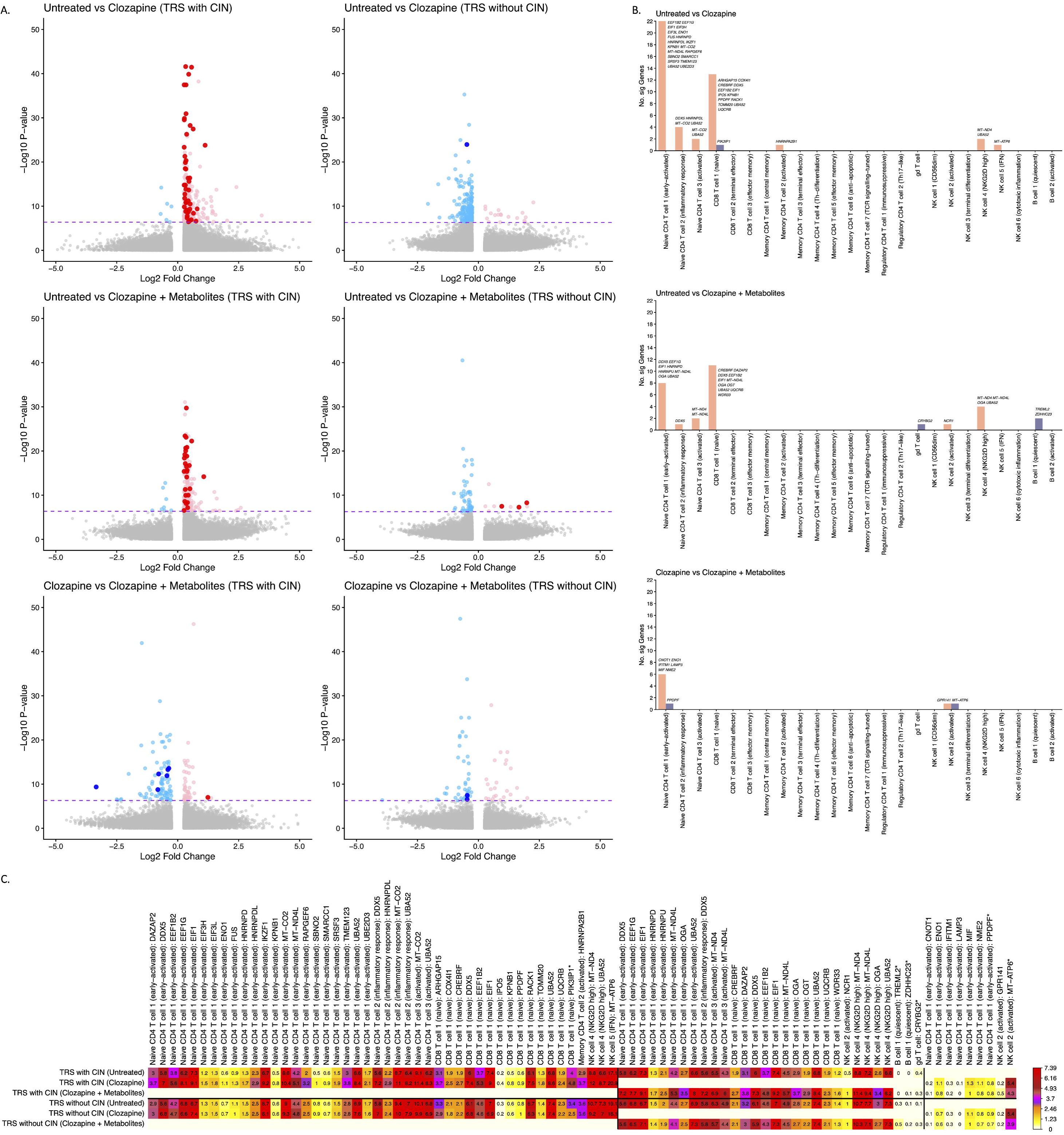
(A) Volcano plots showing differential expression results for three treatment comparisons (untreated vs. clozapine, untreated vs. clozapine + metabolites, and clozapine vs. clozapine + metabolites) stratified by TRS with and without CIN. Significant differentially expressed (DE) genes that passed Bonferroni correction and remained significant in both sensitivity analyses are highlighted. (B) Significant DE genes from panel A, summarised by cluster (number of genes and gene identifiers). (C) Heatmap of average expression for significant DE genes (panel A) across sample and treatment groups.

A similar pattern held for the response to clozapine in combination with its two major metabolites. In TRS with CIN, this combination was associated with cell type-specific upregulation of 27 genes (vs. untreated samples), with a majority in naïve CD4 and CD8 T cells (Table S4). In contrast, only three cell type-specific DE genes were detected in response to clozapine and its metabolites in TRS without CIN (Table S5). Among the 27 genes dysregulated by clozapine + metabolites in TRS with CIN, 15 were also identified in the TRS with CIN group exposed to clozapine alone, including *DDX5*, *EIF1* and *UBA52* that were shared across multiple naïve CD4 and CD8 T cell types. There was no correlation between the log_2_FC of these genes in TRS with CIN and the corresponding estimates in TRS without CIN (*ρ* = -0.05, p-value = 0.80). Additionally, comparing clozapine alone with clozapine + metabolites yielded seven significantly DE genes in TRS with CIN (Tables S6-7). Given comparable statistical power for DE testing across the three group comparisons, the metabolite□specific comparison produced far fewer DE genes than either treatment□versus□untreated contrast, indicating only a mild to moderate incremental effect of the two metabolites on the clozapine□induced transcriptional programs.

### Identification of a cell type-specific gene co-expression module associated with CIN

The application of hdWGCNA to three qualifying clusters (naïve CD4 T cell (early-activated), CD8 T cell (naïve), and naïve CD4 T cell (inflammatory response)) identified 7, 9, and 8 gene co-expression modules, respectively (Figures S12-17). For each of the 24 module-cell type combinations, we performed differential module eigengene (DME) analyses across the seven group and treatment comparisons (as previously described). We identified a single module in naïve CD4 T cells (inflammatory response) with significantly lower eigengene expression in TRS with vs without CIN, after Bonferroni correction (Figure 4A and Table S8). This module comprised 254 genes (Table S9), 42 of which reached Bonferroni significance in the primary negative binomial regression (10 with |log_2_FC| >0.25), with evidence for significant enrichment of DE genes within this module (Fisher’s exact test: odds ratio (OR) = 34.29; 95% confidence interval (CI) = 22.1 – 52.8; p-value = 2.64×10^-42^). Transcription factor (TF) regulatory network analyses of this module identified one candidate TF (SREBF1). Negative regulons for SREBF1 were significantly inversely associated (after Bonferroni correction) with TRS with versus without CIN (Figure 4B and Table S10).

**Figure 4.**
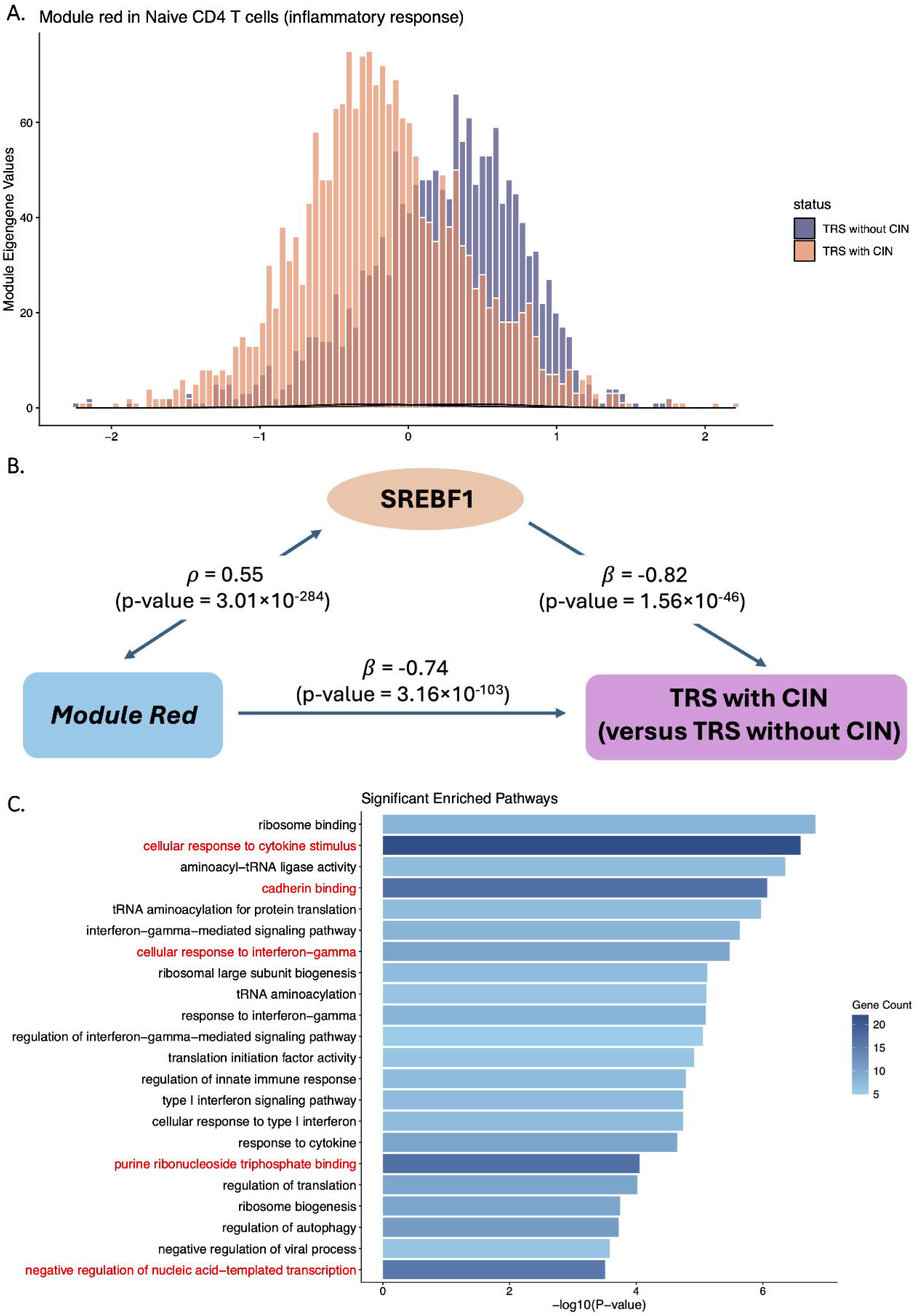
(A) Distribution of the module eigengene (inverse rank based transformed) for the red module in naïve CD4 T cell (inflammatory response) in TRS with and without CIN. (B) Candidate transcription factor (SREBF1) and its association with TRS with CIN and the red module in naïve CD4 T cell (inflammatory response). (C) Significantly enriched pathways (enrichR) underlying the red module in naïve CD4 T cell (inflammatory response). Pathways shown in red were further identified by SCPA.

Pathway enrichment analyses using enrichR identified 22 Bonferroni-significant pathways in this module (Figure 4C and Table S11), of which five also reached Bonferroni significance using SCPA, highlighting cytokine and interferon-gamma responses, transcriptional repression, and cadherin and nucleotide binding. No pathways reached Bonferroni significance for known targets of SREBF1, likely reflecting limited statistical power.

### Differential cell-cell communication patterns associated with CIN in TRS and response to clozapine and its metabolites

Finally, cell-cell communication analysis revealed seven ligand-receptor (L-R) pairs. Five of these (“*CD99*–*CD99*”, “*CD99*–*PILRB*”, “*CLEC2B*–*KLRB1*”, “*CLEC2D*–*KLRB1*”, “*ICAM2*– (*ITGAL*+*ITGB2*)”) showed significantly different communication probabilities between TRS with vs without CIN (Figure 5A and Table S12). All five L-R pairs mediated communication among CD4 T cells, CD8 T cells, and NK cells. Specifically, “*CLEC2B*–*KLRB1*” and “*CLEC2D*–*KLRB1*” signalling originated from naïve CD4/CD8 T cells and NK cells (NKG2D high) and converged on NK cells (terminal differentiation), whereas memory CD4 T cells (Th-differentiation) and NK cells (terminal differentiation) were the main sources of “*CD99*–*CD99*”, “*CD99*–*PILRB*”, and “*ICAM2*–(*ITGAL*+*ITGB2*)” signalling.

**Figure 5.**
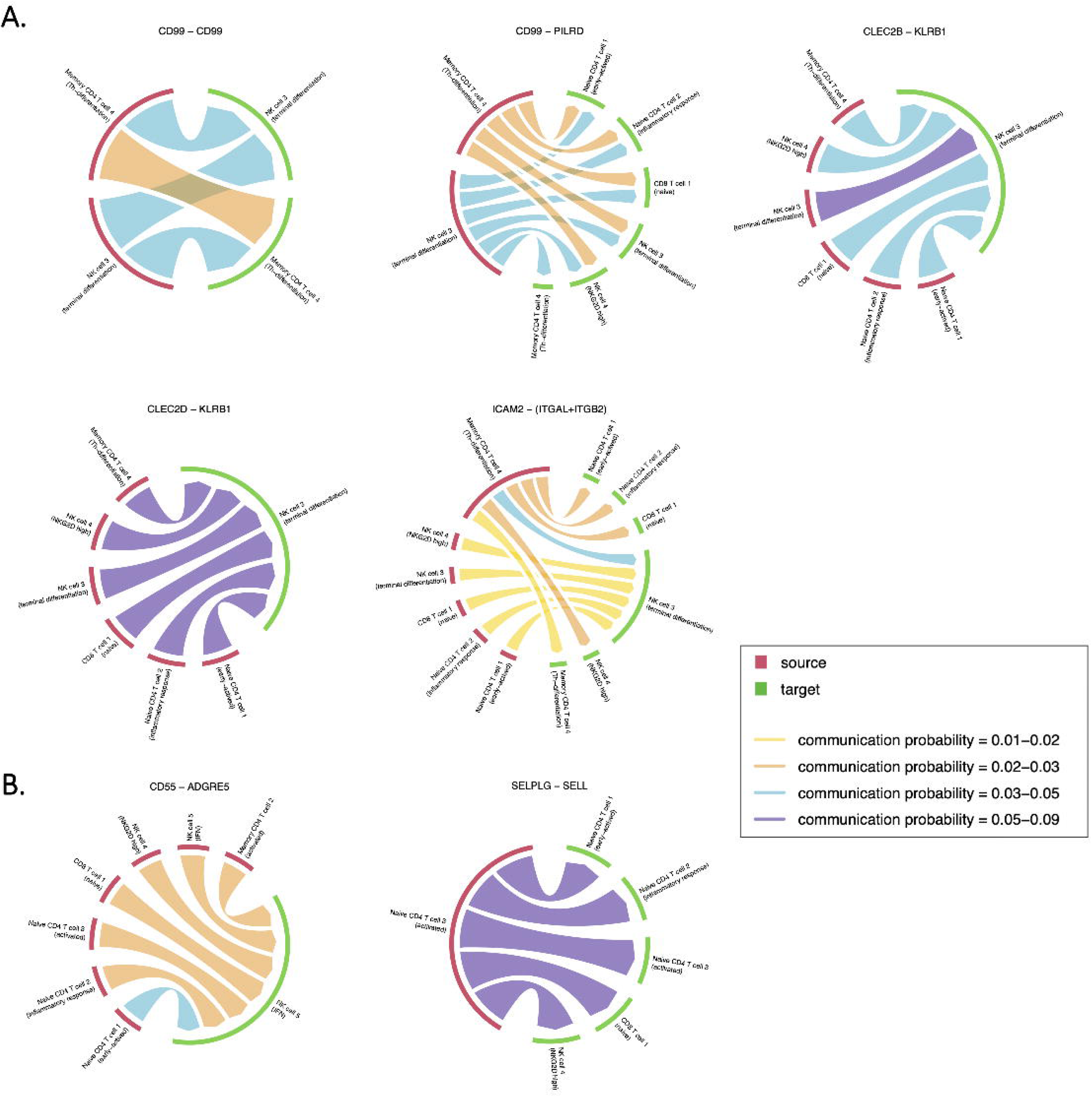
(A) Ligand-receptor (L-R) pair-specific cell-cell communication networks that differed significantly between TRS with vs without CIN. (B) L-R pair-specific cell-cell communication networks significantly associated with response to clozapine (and its metabolites; vs. untreated) in TRS with CIN but not in TRS without CIN.

The two remaining L-R pairs (“*CD55*–*ADGRE5*” for clozapine alone and “*SELPLG*–*SELL*” for clozapine + metabolites) were significantly associated with clozapine response (vs. untreated) in TRS with CIN but not in TRS without CIN (Figure 5B and Table S12). “*CD55*– *ADGRE5*” participated in intercellular signalling from multiple CD4 T cells, CD8 T cells and NK cells to NK cells (IFN), whereas “*SELPLG*–*SELL*” signalling originated from naïve CD4 T cells (activated) to naïve CD4 T cells, CD8 T cells (naïve) and NK cells (NKG2D high). Conversely, no L-R pairs showed significant changes that were specific to TRS without CIN, and no significant L-R pairs were identified specific to clozapine + metabolites (vs. clozapine), consistent with our earlier analyses.

## Discussion

To the best of our knowledge, this is the first human single nucleus RNA-seq study to characterise PBMC-based cellular transcriptomic changes associated with CIN and response to clozapine and its two major metabolites in TRS with and without CIN. We identified a set of cell type-specific differentially expressed (DE) genes associated with CIN status, as well as DE genes that respond to clozapine (and its metabolites) in TRS with CIN, but not TRS without CIN, most of which have not been reported in prior GWAS of CIN. Co-expression analyses provided convergent evidence for a central role of a naïve CD4 T cell (inflammatory response)-specific module in TRS with CIN, potentially regulated by the SREBF1. Pathway analysis implicated cytokine and interferon-gamma responses, transcriptional repression, and cadherin and nucleotide binding underlying this co-expression module. Additionally, *CD99*-, *CLEC*-, and *ICAM*-associated L-R pairs were implicated in association with TRS with CIN, while *SELPLG*- and *ADGRE*-associated L-R pairs were linked to clozapine response in TRS with CIN only, across multiple cell type interactions, suggesting altered intercellular communication in CIN and following clozapine exposure.

The key contribution of this study is the generation of the first single nucleus transcriptomic landscape of PBMCs for response to clozapine and its two major metabolites in TRS with and without CIN, resolving 26 distinct immune cell populations and a list of 57 cell type-specific DE genes associated with CIN status and exposure to clozapine and/or its metabolites. Among the CIN-associated genes, *FAM118A* and *IL32* were notable for being significantly differentially expressed across multiple immune cell types. *FAM118A* encodes a noncanonical human sirtuin protein that depletes nicotinamide adenine dinucleotide (NAD^+^) when overexpressed (41), which has the potential to impact NAD-dependent granulopoiesis (42). We found that *FAM118A* was consistently upregulated in naïve CD4 and CD8 T cells and NK cells in TRS with CIN, suggesting that increased *FAM118A* expression may disrupt NAD-dependent signals that support neutrophil production. *IL32* encodes a pro-inflammatory cytokine that induces TNF-α and IL-8 (43, 44), and binds proteinase 3, a serine protease abundantly expressed in neutrophils (45), suggesting a potential link between lymphocyte activation and neutrophil function.

Beyond these CIN-associated genes, we identified a distinct set of cell type-specific DE genes associated with exposure to clozapine and/or clozapine + metabolites in TRS donors with CIN but not in TRS without CIN. Two of the identified genes, *DDX5* and *UBA52* are of particular interest. *DDX5* acts as a transcriptional corepressor that restricts the suppressive activity of RORγt□regulatory T cells by reducing IL-10 production (46). *DDX5* was upregulated in naïve CD4 and CD8 T cells following clozapine exposure, which may reflect altered T cell activation or regulation, consistent with prior evidence that clozapine modulates immune signalling (47). The *UBA52* gene encodes a precursor comprising ubiquitin fused to the 60S ribosomal protein eL40, linking protein homeostasis to translation (48, 49). In our study, *UBA52* was upregulated across naïve CD4 T, CD8 T and NK cells after exposure to both clozapine and clozapine plus metabolites, suggesting a consistent clozapine-associated transcriptional response in TRS with CIN, rather than a metabolite-dependent effect. *UBA52* was also associated with CIN status when comparing TRS with CIN to TRS without CIN, but in different cell types and with inconsistent direction of expression change, suggesting context-dependent regulation rather than serving as a fixed marker of neutropenia.

Interestingly, querying the 57 significant cell type-specific DE genes in our analysis with the Drug Gene Interaction Database (DGIdb) (50) revealed 43 approved drug-gene interactions (Table S13), including three curated in ChEMBL (*NDUFA12*, *MT-ND4*, *MT-ND4L*) that all pointed to metformin hydrochloride. Metformin has been reported to mitigate clozapine-induced weighted gain, BMI increase and associated metabolic disturbances (51–53). Our findings suggest that metformin may warrant investigation for repurposing in CIN, although further validation is needed, as all identified drug-gene interactions were supported by only a single source.

More broadly, although no genes overlapped between the CIN-associated and clozapine-related gene sets at Bonferroni significance in our analyses, we observed significant correlations between the two when the full gene lists were compared (Figure S18). CIN-associated gene expression changes were weakly positively correlated with clozapine response in TRS with CIN (*ρ* = 0.05, p-value = 1.72×10^-32^), and negatively correlated with clozapine response in TRS without CIN (*ρ* = -0.22, p-value < 1×10^-300^). The direction of these correlations is consistent with our expectations: CIN-associated transcriptional alterations should align with clozapine-related changes in TRS with CIN, whereas the relationship is anticipated to be reversed in TRS without CIN. The same pattern held for analyses involving response to clozapine + metabolites, whereas it was weaker for the direct comparison of clozapine alone with clozapine + metabolites (TRS with CIN: =*ρ* 0.03, p-value = 1.30×10^-11^; TRS without CIN:*ρ* = 0.06, p-value = 1.80×10^-34^). This suggests that a shared transcriptional signal exists but is too subtle for individual genes to pass stringent multiple-testing correction in either analysis. Differences in study design and statistical power, together with cell type-specific effects, may further obscure this overlap. Validation in larger cohorts will be needed to confirm and characterise this shared signal.

Furthermore, we identified a module in naïve CD4 T cells (inflammatory response) that was associated with CIN status and potentially regulated by the transcription factor SREBF1. The top hub genes in this module form a prominent functional group comprising genes involved in translation initiation and elongation, including *EEF1G*, *EEF2*, and *EEF1B2*. The eEF1B complex (*EEF1B2*, *EEF1G*) regenerates eEF1A-GTP to sustain aminoacyl-tRNA delivery during elongation (54), and *EEF2* drives ribosomal translocation that is regulated by mTORC1 signalling (55). The coordinated upregulation of these genes is consistent with the translational preparedness of naïve T cells, which maintain idling ribosomes and untranslated mRNAs that support rapid responses upon activation (56, 57). SREBP1 activity is likewise regulated downstream of mTORC1 (58), and is required for the metabolic reprogramming that supports T cell expansion (59), including through a TCR-mTORC1-SREBP1 axis in CD4 T cells (60). The co-occurrence of elongation genes and a putative SREBF1-regulated module therefore suggests a shared mTORC1-responsive growth programme, in which translational capacity and lipid biosynthesis are coordinated.

Turning to intercellular communication, we identified five ligand-receptor pairs (Figure 5A) associated with CIN (vs. no CIN). Notably, both the *CD99*-associated L-R pairs (“*CD99*– *CD99*” and “*CD99*–*PILRB*”) and the ICAM-associated pairs (“*ICAM2*–(*ITGAL*+*ITGB2*)”) are actively involved in leukocyte adhesion and trans-endothelial migration. *CD99* blockade impairs lymphocyte and neutrophil extravasation *in vivo* (61, 62). Similarly, binding of LFA-1 (*ITGAL*+*ITGB2*) to endothelial *ICAM* molecules mediates firm neutrophil adhesion (63). These pathways may be relevant to CIN, as some cases of CIN have been hypothesised to result from enhanced neutrophil margination rather than impaired bone marrow production (64).

Separately, we identified two L-R pairs (Figure 5B; “*CD55*–*ADGRE5*”, “*SELPLG*–*SELL*”) associated with clozapine and/or clozapine + metabolites (vs. untreated). The “*CD55*– *ADGRE5*” interaction was predicted from memory CD4 T cells to NK cells under clozapine exposure. *CD97*, encoded by *ADGRE5*, is broadly expressed on leukocytes and interacts with *CD55* to regulate immune cell adhesion and migration (65), and this interaction has been functionally demonstrated to regulate granulocyte homeostasis in myeloid cells (66), providing a potentially relevant but indirect link to clozapine. In contrast, “*SELPLG*–*SELL*” contributes to leukocyte tethering and rolling during the early stages of leukocyte trafficking and extravasation (67, 68). Together, these findings suggest altered immune-cell adhesion and trafficking programmes in TRS with CIN.

Our findings have important clinical implications. Despite its superior efficacy in TRS, clozapine remains underutilised, partly due to concerns regarding serious adverse effects such as CIN (69). Current management of CIN typically involves clozapine withdrawal, with potential for cautious rechallenge if alternative contributing factors are identified and addressed. Granulocyte colony-stimulating factor and lithium have shown promise in supporting neutrophil counts during clozapine rechallenge (70). However, a major clinical challenge remains the inability to predict which individuals are susceptible to CIN or its recurrence. Our findings support an immune-mediated mechanism underlying CIN and identify candidate cell type-specific genes and pathways that may play a role in susceptibility. Furthermore, by characterising cell-specific responses, our study identifies candidate mechanisms that may inform the development of targeted therapeutic strategies to prevent CIN before clinically significant neutropenia develops. Such advances could facilitate the safe continuation or reintroduction of clozapine in individuals with TRS, for whom effective treatment alternatives remain limited, with potentially life-saving implications.

Our findings should be interpreted in the context of several limitations. First, we used the 10X Genomics Fixed RNA Profiling platform for snRNA-seq, which excludes hypervariable genes in the HLA region, a well-known genetic locus associated with clozapine response; this may have constrained our ability to detect all relevant associations. Second, our study including only three TRS donors with CIN and three without, primarily due to the low prevalence of CIN and the recruitment challenges inherent to a severely ill patient cohort. Consequently, the study lacks the statistical power to detect cell type-specific DE genes, co-expression modules, and pathways with small effect sizes, and we cannot entirely account for the influence of confounding variables such as age, sex and smoking. Consistent with this limitation, intersecting the 57 significant cell type-specific DE genes identified in our study with MAGMA gene-based results from GWAS of CIN (17) yielded no overlap after multiple-testing correction (Supplementary Methods, Table S14). Larger cohorts will be necessary to validate and expand upon these preliminary findings. Third, we only focused on the two major clozapine-related metabolites. Potential effects of other bioactivated or downstream metabolites of clozapine remain to be defined and warrant systematic evaluation. Finally, the lack of an independent replication cohort necessitates caution in the interpretation of the results.

In summary, this study provides the first single nucleus transcriptomic characterisation of immune cell type-specific responses to clozapine and its two major metabolites in TRS with and without CIN. We identified a set of cell type-specific DE genes, predominantly in naïve CD4 and CD8 T cells, a naïve CD4 T cell-specific co-expression module, and five intercellular L-R pairs that was associated with TRS with CIN compared to TRS without CIN. We also identified distinct cell type-specific DE genes and L-R pairs involved in response to clozapine in TRS with CIN but not TRS without CIN. While several DE genes, such as *FAM118A* and *DDX5*, provide biologically plausible links to neutropenia-relevant mechanisms, these associations remain exploratory pending validation. Future studies should replicate these findings in larger, independent cohorts, including prospective sampling before clozapine initiation and comparison against other immune-mediated drug-induced agranulocytosis, to establish both temporality and specificity. Functional evaluation of the implicated pathways in naïve CD4 T cells co-cultured with metabolite-pulsed autologous antigen-presenting cells would test whether the observed transcriptional state modifies the activation threshold to clozapine-derived antigen. Ultimately, with sufficient validation, these findings may contribute to the long-term development of a clinical tool for stratifying CIN risk in patients with TRS, potentially informing more tailored clinical monitoring.

## Data Availability

All data produced in the present study are available upon reasonable request to the authors.

## Acknowledgments

The authors acknowledge the Translational Research Institute (TRI) the Leading Innovations through New Collaborations (LINC) grant for supporting the work. Y.Y and J.G acknowledge support from Mater Foundation and Mater Research. J.G acknowledges support from the Australian National Health and Medical Research Council (NHMRC) GNT2018431. D.S is supported by a UQ Amplify Professorial Research Fellowship (2026) and NHMRC GNT2051821. Claude (Anthropic) and Copilot (GPT-5) were used to improve the language, clarity, and grammar of the manuscript. All authors subsequently reviewed and edited the text.

## Author contributions

Y.Y, T.E, D.S and J.G designed the project. T.E carried out sample recruitment and blood collection; T.E, N.J, M.O.P and S.N performed experiments; Y.Y led the data analysis, with input from JG and support from P.K and C.S on the interpretation of immune signatures. D.S and J.G provided supervision; Y.Y and T.E wrote the manuscript. All authors reviewed and approved the final version of the manuscript.

## References

1. McCutcheon RA, Reis Marques T, Howes OD (2020): Schizophrenia-An Overview. JAMA Psychiatry. 77:201–210.

2. Howes OD, McCutcheon R, Agid O, de Bartolomeis A, van Beveren NJ, Birnbaum ML, et al. (2017): Treatment-Resistant Schizophrenia: Treatment Response and Resistance in Psychosis (TRRIP) Working Group Consensus Guidelines on Diagnosis and Terminology. Am J Psychiatry. 174:216–229.

3. Siskind D, Orr S, Sinha S, Yu O, Brijball B, Warren N, et al. (2022): Rates of treatment-resistant schizophrenia from first-episode cohorts: systematic review and meta-analysis. Br J Psychiatry. 220:115–120.

4. Siskind D, Siskind V, Kisely S (2017): Clozapine Response Rates among People with Treatment-Resistant Schizophrenia: Data from a Systematic Review and Meta-Analysis. Can J Psychiatry. 62:772–777.

5. Land R, Siskind D, McArdle P, Kisely S, Winckel K, Hollingworth SA (2017): The impact of clozapine on hospital use: a systematic review and meta-analysis. Acta Psychiatr Scand. 135:296–309.

6. Tiihonen J, Lonnqvist J, Wahlbeck K, Klaukka T, Niskanen L, Tanskanen A, et al. (2009): 11-year follow-up of mortality in patients with schizophrenia: a population-based cohort study (FIN11 study). Lancet. 374:620–627.

7. Meltzer HY, Alphs L, Green AI, Altamura AC, Anand R, Bertoldi A, et al. (2003): Clozapine treatment for suicidality in schizophrenia: International Suicide Prevention Trial (InterSePT). Arch Gen Psychiatry. 60:82–91.

8. Vermeulen JM, van Rooijen G, van de Kerkhof MPJ, Sutterland AL, Correll CU, de Haan L (2019): Clozapine and Long-Term Mortality Risk in Patients With Schizophrenia: A Systematic Review and Meta-analysis of Studies Lasting 1.1-12.5 Years. Schizophr Bull. 45:315–329.

9. Siskind D, McCartney L, Goldschlager R, Kisely S (2016): Clozapine v. first- and second-generation antipsychotics in treatment-refractory schizophrenia: systematic review and meta-analysis. Br J Psychiatry. 209:385–392.

10. Forrester T, Siskind D, Winckel K, Wheeler A, Hollingworth S (2015): Increasing Clozapine Dispensing Trends in Queensland, Australia 2004-2013. Pharmacopsychiatry. 48:164–169.

11. Myles N, Myles H, Xia S, Large M, Kisely S, Galletly C, et al. (2018): Meta-analysis examining the epidemiology of clozapine-associated neutropenia. Acta Psychiatr Scand. 138:101–109.

12. Northwood K, Myles N, Clark SR, Every-Palmer S, Myles H, Kisely S, et al. (2024): Evaluating the epidemiology of clozapine-associated neutropenia among people on clozapine across Australia and Aotearoa New Zealand: a retrospective cohort study. Lancet Psychiatry. 11:27–35.

13. Crilly J (2007): The history of clozapine and its emergence in the US market: a review and analysis. Hist Psychiatry. 18:39–60.

14. Siskind D, Northwood K, Pillinger T, Chan S, Correll C, Cotes RO, et al. (2026): Absolute neutrophil count and adverse drug reaction monitoring during clozapine treatment: consensus guidelines from a global Delphi panel. Lancet Psychiatry. 13:77–86.

15. U.S. Food and Drug Administration (2025): FDA removes risk evaluation and mitigation strategy (REMS) program for the antipsychotic drug Clozapine.

16. Rubio JM, Kane JM, Tanskanen A, Tiihonen J, Taipale H (2024): Long-term persistence of the risk of agranulocytosis with clozapine compared with other antipsychotics: a nationwide cohort and case-control study in Finland. Lancet Psychiatry. 11:443–450.

17. Legge SE, Hamshere ML, Ripke S, Pardinas AF, Goldstein JI, Rees E, et al. (2017): Genome-wide common and rare variant analysis provides novel insights into clozapine-associated neutropenia. Mol Psychiatry. 22:1502–1508.

18. Chen J, Yang P, Zhang Q, Chen R, Wang P, Liu B, et al. (2021): Genetic risk of clozapine-induced leukopenia and neutropenia: a genome-wide association study. Transl Psychiatry. 11:343.

19. Legge SE, Walters JT (2019): Genetics of clozapine-associated neutropenia: recent advances, challenges and future perspective. Pharmacogenomics. 20:279–290.

20. Lin BD, Pinzon-Espinosa J, Blouzard E, van der Horst MZ, Okhuijsen-Pfeifer C, van Eijk KR, et al. (2023): Associations Between Polygenic Risk Score Loading, Psychosis Liability, and Clozapine Use Among Individuals With Schizophrenia. JAMA Psychiatry. 80:181–185.

21. Regen F, Herzog I, Hahn E, Ruehl C, Le Bret N, Dettling M, et al. (2017): Clozapine-induced agranulocytosis: Evidence for an immune-mediated mechanism from a patient-specific in-vitro approach. Toxicol Appl Pharmacol. 316:10–16.

22. Liu ZC, Uetrecht JP (1995): Clozapine is oxidized by activated human neutrophils to a reactive nitrenium ion that irreversibly binds to the cells. J Pharmacol Exp Ther. 275:1476–1483.

23. Wicinski M, Weclewicz MM, Mietkiewicz M, Malinowski B, Grzesk E, Klonowska J (2017): Potential Mechanisms of Hematological Adverse Drug Reactions in Patients Receiving Clozapine in Combination With Proton Pump Inhibitors. J Psychiatr Pract. 23:114–120.

24. Gerson SL, Arce C, Meltzer HY (1994): N-desmethylclozapine: a clozapine metabolite that suppresses haemopoiesis. Br J Haematol. 86:555–561.

25. Goossens J, Morrens M, Coppens V (2021): The Potential Use of Peripheral Blood Mononuclear Cells as Biomarkers for Treatment Response and Outcome Prediction in Psychiatry: A Systematic Review. Mol Diagn Ther. 25:283–299.

26. Aravagiri M, Marder SR (2001): Simultaneous determination of clozapine and its N-desmethyl and N-oxide metabolites in plasma by liquid chromatography/electrospray tandem mass spectrometry and its application to plasma level monitoring in schizophrenic patients. J Pharm Biomed Anal. 26:301–311.

27. Correll CU, Martin A, Patel C, Benson C, Goulding R, Kern-Sliwa J, et al. (2022): Systematic literature review of schizophrenia clinical practice guidelines on acute and maintenance management with antipsychotics. Schizophrenia (Heidelb). 8:5.

28. Zheng GX, Terry JM, Belgrader P, Ryvkin P, Bent ZW, Wilson R, et al. (2017): Massively parallel digital transcriptional profiling of single cells. Nat Commun. 8:14049.

29. Wolock SL, Lopez R, Klein AM (2019): Scrublet: Computational Identification of Cell Doublets in Single-Cell Transcriptomic Data. Cell Syst. 8:281–291 e289.

30. Hafemeister C, Satija R (2019): Normalization and variance stabilization of single-cell RNA-seq data using regularized negative binomial regression. Genome Biol. 20:296.

31. Hao Y, Hao S, Andersen-Nissen E, Mauck WM, 3rd, Zheng S, Butler A, et al. (2021): Integrated analysis of multimodal single-cell data. Cell. 184:3573–3587 e3529.

32. Aran D, Looney AP, Liu L, Wu E, Fong V, Hsu A, et al. (2019): Reference-based analysis of lung single-cell sequencing reveals a transitional profibrotic macrophage. Nat Immunol. 20:163–172.

33. de Cevins C, Delage L, Batignes M, Riller Q, Luka M, Remaury A, et al. (2023): Single-cell RNA-sequencing of PBMCs from SAVI patients reveals disease-associated monocytes with elevated integrated stress response. Cell Rep Med. 4:101333.

34. Hu C, Li T, Xu Y, Zhang X, Li F, Bai J, et al. (2023): CellMarker 2.0: an updated database of manually curated cell markers in human/mouse and web tools based on scRNA-seq data. Nucleic Acids Res. 51:D870–D876.

35. Bates D, Mächler M, Bolker B, Walker S (2015): Fitting Linear Mixed-Effects Models Using lme4. Journal of Statistical Software. 67:1–48.

36. Morabito S, Reese F, Rahimzadeh N, Miyoshi E, Swarup V (2023): hdWGCNA identifies co-expression networks in high-dimensional transcriptomics data. Cell Rep Methods. 3:100498.

37. Rauluseviciute I, Riudavets-Puig R, Blanc-Mathieu R, Castro-Mondragon JA, Ferenc K, Kumar V, et al. (2024): JASPAR 2024: 20th anniversary of the open-access database of transcription factor binding profiles. Nucleic Acids Res. 52:D174–D182.

38. Kuleshov MV, Jones MR, Rouillard AD, Fernandez NF, Duan Q, Wang Z, et al. (2016): Enrichr: a comprehensive gene set enrichment analysis web server 2016 update. Nucleic Acids Res. 44:W90–97.

39. Bibby JA, Agarwal D, Freiwald T, Kunz N, Merle NS, West EE, et al. (2022): Systematic single-cell pathway analysis to characterize early T cell activation. Cell Rep. 41:111697.

40. Jin S, Plikus MV, Nie Q (2025): CellChat for systematic analysis of cell-cell communication from single-cell transcriptomics. Nat Protoc. 20:180–219.

41. Baretic D, Missoury S, Patel K, Martinez M, Coste F, Zhu K, et al. (2025): Filament formation and NAD processing by noncanonical human FAM118 sirtuins. Nat Struct Mol Biol. 32:2526–2541.

42. Skokowa J, Lan D, Thakur BK, Wang F, Gupta K, Cario G, et al. (2009): NAMPT is essential for the G-CSF-induced myeloid differentiation via a NAD(+)-sirtuin-1-dependent pathway. Nat Med. 15:151–158.

43. Dinarello CA, Kim SH (2006): IL-32, a novel cytokine with a possible role in disease. Ann Rheum Dis. 65 Suppl 3:iii61–64.

44. Kim SH, Han SY, Azam T, Yoon DY, Dinarello CA (2005): Interleukin-32: a cytokine and inducer of TNFalpha. Immunity. 22:131–142.

45. Novick D, Rubinstein M, Azam T, Rabinkov A, Dinarello CA, Kim SH (2006): Proteinase 3 is an IL-32 binding protein. Proc Natl Acad Sci U S A. 103:3316–3321.

46. Ma S, Yang Q, Chen N, Zheng A, Abbasi N, Wang G, et al. (2023): RNA binding protein DDX5 restricts RORgammat(+) T(reg) suppressor function to promote intestine inflammation. Sci Adv. 9:eadd6165.

47. Chen ML, Tsai TC, Lin YY, Tsai YM, Wang LK, Lee MC, et al. (2011): Antipsychotic drugs suppress the AKT/NF-kappaB pathway and regulate the differentiation of T-cell subsets. Immunol Lett. 140:81–91.

48. Webb GC, Baker RT, Coggan M, Board PG (1994): Localization of the human UBA52 ubiquitin fusion gene to chromosome band 19p13.1-p12. Genomics. 19:567–569.

49. Kobayashi M, Oshima S, Maeyashiki C, Nibe Y, Otsubo K, Matsuzawa Y, et al. (2016): The ubiquitin hybrid gene UBA52 regulates ubiquitination of ribosome and sustains embryonic development. Sci Rep. 6:36780.

50. Freshour SL, Kiwala S, Cotto KC, Coffman AC, McMichael JF, Song JJ, et al. (2021): Integration of the Drug-Gene Interaction Database (DGIdb 4.0) with open crowdsource efforts. Nucleic Acids Res. 49:D1144–D1151.

51. Chiu CC, Lu ML, Huang MC, Chen PY, Lin YK, Lin SK, et al. (2016): Effects of Low Dose Metformin on Metabolic Traits in Clozapine-Treated Schizophrenia Patients: An Exploratory Twelve-Week Randomized, Double-Blind, Placebo-Controlled Study. PLoS One. 11:e0168347.

52. Liu Z, Zheng W, Gao S, Qin Z, Li G, Ning Y (2015): Metformin for treatment of clozapine-induced weight gain in adult patients with schizophrenia: a meta-analysis. Shanghai Arch Psychiatry. 27:331–340.

53. Stogios N, Maksyutynska K, Navagnanavel J, Sanches M, Powell V, Gerretsen P, et al. (2022): Metformin for the prevention of clozapine-induced weight gain: A retrospective naturalistic cohort study. Acta Psychiatr Scand. 146:190–200.

54. Le Sourd F, Boulben S, Le Bouffant R, Cormier P, Morales J, Belle R, et al. (2006): eEF1B: At the dawn of the 21st century. Biochim Biophys Acta. 1759:13–31.

55. Proud CG (2015): Regulation and roles of elongation factor 2 kinase. Biochem Soc Trans. 43:328–332.

56. Wolf T, Jin W, Zoppi G, Vogel IA, Akhmedov M, Bleck CKE, et al. (2020): Dynamics in protein translation sustaining T cell preparedness. Nat Immunol. 21:927–937.

57. Ricciardi S, Manfrini N, Alfieri R, Calamita P, Crosti MC, Gallo S, et al. (2018): The Translational Machinery of Human CD4(+) T Cells Is Poised for Activation and Controls the Switch from Quiescence to Metabolic Remodeling. Cell Metab. 28:895–906 e895.

58. Duvel K, Yecies JL, Menon S, Raman P, Lipovsky AI, Souza AL, et al. (2010): Activation of a metabolic gene regulatory network downstream of mTOR complex 1. Mol Cell. 39:171–183.

59. Kidani Y, Elsaesser H, Hock MB, Vergnes L, Williams KJ, Argus JP, et al. (2013): Sterol regulatory element-binding proteins are essential for the metabolic programming of effector T cells and adaptive immunity. Nat Immunol. 14:489–499.

60. Angela M, Endo Y, Asou HK, Yamamoto T, Tumes DJ, Tokuyama H, et al. (2016): Fatty acid metabolic reprogramming via mTOR-mediated inductions of PPARgamma directs early activation of T cells. Nat Commun. 7:13683.

61. Li YT, Goswami D, Follmer M, Artz A, Pacheco-Blanco M, Vestweber D (2019): Blood flow guides sequential support of neutrophil arrest and diapedesis by PILR-beta1 and PILR-alpha. Elife. 8.

62. Goswami D, Marz S, Li YT, Artz A, Schafer K, Seelige R, et al. (2017): Endothelial CD99 supports arrest of mouse neutrophils in venules and binds to neutrophil PILRs. Blood. 129:1811–1822.

63. Smith CW, Marlin SD, Rothlein R, Toman C, Anderson DC (1989): Cooperative interactions of LFA-1 and Mac-1 with intercellular adhesion molecule-1 in facilitating adherence and transendothelial migration of human neutrophils in vitro. J Clin Invest. 83:2008–2017.

64. Miller BJ, Ghani N, Kota V (2026): Clozapine-Induced “Pseudo-Agranulocytosis”: Bone Marrow Suppression or Increased Neutrophil Margination? Schizophr Bull. 52.

65. Hamann J, Vogel B, van Schijndel GM, van Lier RA (1996): The seven-span transmembrane receptor CD97 has a cellular ligand (CD55, DAF). J Exp Med. 184:1185–1189.

66. Veninga H, Hoek RM, de Vos AF, de Bruin AM, An FQ, van der Poll T, et al. (2011): A novel role for CD55 in granulocyte homeostasis and anti-bacterial host defense. PLoS One. 6:e24431.

67. McEver RP (2015): Selectins: initiators of leucocyte adhesion and signalling at the vascular wall. Cardiovasc Res. 107:331–339.

68. Stadtmann A, Germena G, Block H, Boras M, Rossaint J, Sundd P, et al. (2013): The PSGL-1-L-selectin signaling complex regulates neutrophil adhesion under flow. J Exp Med. 210:2171–2180.

69. Verdoux H, Quiles C, Bachmann CJ, Siskind D (2018): Prescriber and institutional barriers and facilitators of clozapine use: A systematic review. Schizophr Res. 201:10–19.

70. Myles N, Myles H, Clark SR, Bird R, Siskind D (2017): Use of granulocyte-colony stimulating factor to prevent recurrent clozapine-induced neutropenia on drug rechallenge: A systematic review of the literature and clinical recommendations. Aust N Z J Psychiatry. 51:980–989.

