## Supplementary Methods and Figures for "Unravelling the mechanisms of clozapine-induced neutropenia in treatment-resistant schizophrenia using cellular genomics"

**MAGMA gene-based association analysis**

For each significant cell type-specific DE gene, we further evaluated whether it showed evidence of genome-wide significant association in an independent genome-wide association study (GWAS) of CIN (66 cases; 5,583 controls) (1). Gene-based association analysis was performed via Multi-marker Analysis of GenoMic Annotation (MAGMA) (2), using the European-ancestry 1000 Genomes reference panel to account for linkage disequilibrium (LD) structure. Single nucleotide polymorphisms (SNPs) were mapped to candidate genes using three upstream and downstream window definitions relative to each gene boundary: (i) a Hi-C coupled MAGMA-informed window (35 kb upstream, 10 kb downstream); (ii) a ±100 kb proximal regulatory window; and (iii) a ±250 kb distal regulatory window.

**References**

1.	Legge SE, Hamshere ML, Ripke S, Pardinas AF, Goldstein JI, Rees E, et al. (2017): Genome-wide common and rare variant analysis provides novel insights into clozapine-associated neutropenia. *Mol Psychiatry*. 22:1502-1508.

2.	de Leeuw CA, Mooij JM, Heskes T, Posthuma D (2015): MAGMA: generalized gene-set analysis of GWAS data. *PLoS Comput Biol*. 11:e1004219.

**Supplementary Figures**

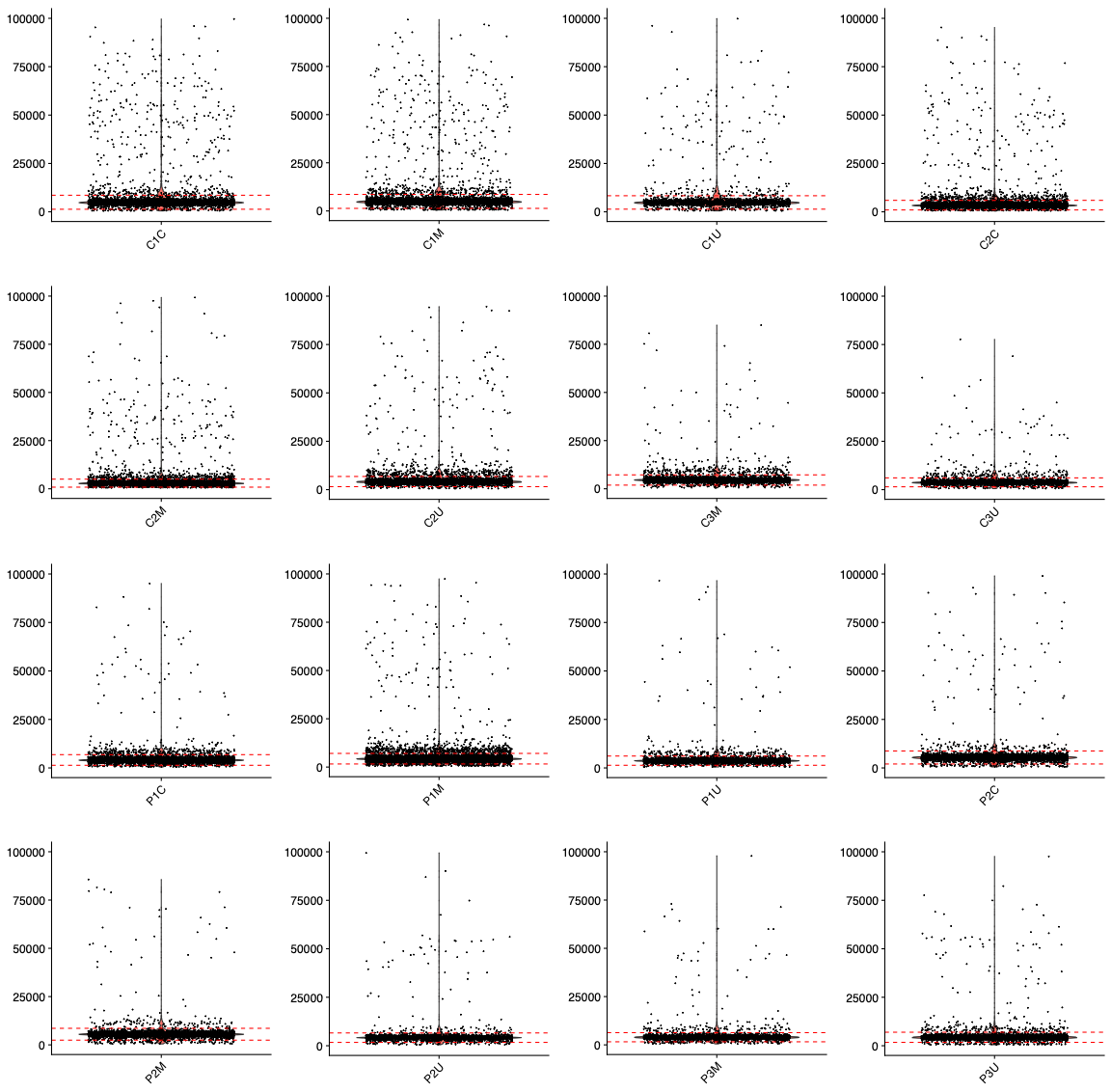

**Figure S1.** Distribution of the unique molecular identifier (UMI) count per gene per cell for each individual sample. The red dotted line indicates the cutoff, defined as the median UMI count per cell ± 3 median absolute deviations (MAD). Sample IDs use a three-character code: the first character indicates sample status (“P” for TRS with CIN, “C” for TRS without CIN), the second is a numeric sample identifier, and the third indicates treatment condition (“U” = untreated, “C” = clozapine, “M” = clozapine + metabolites).

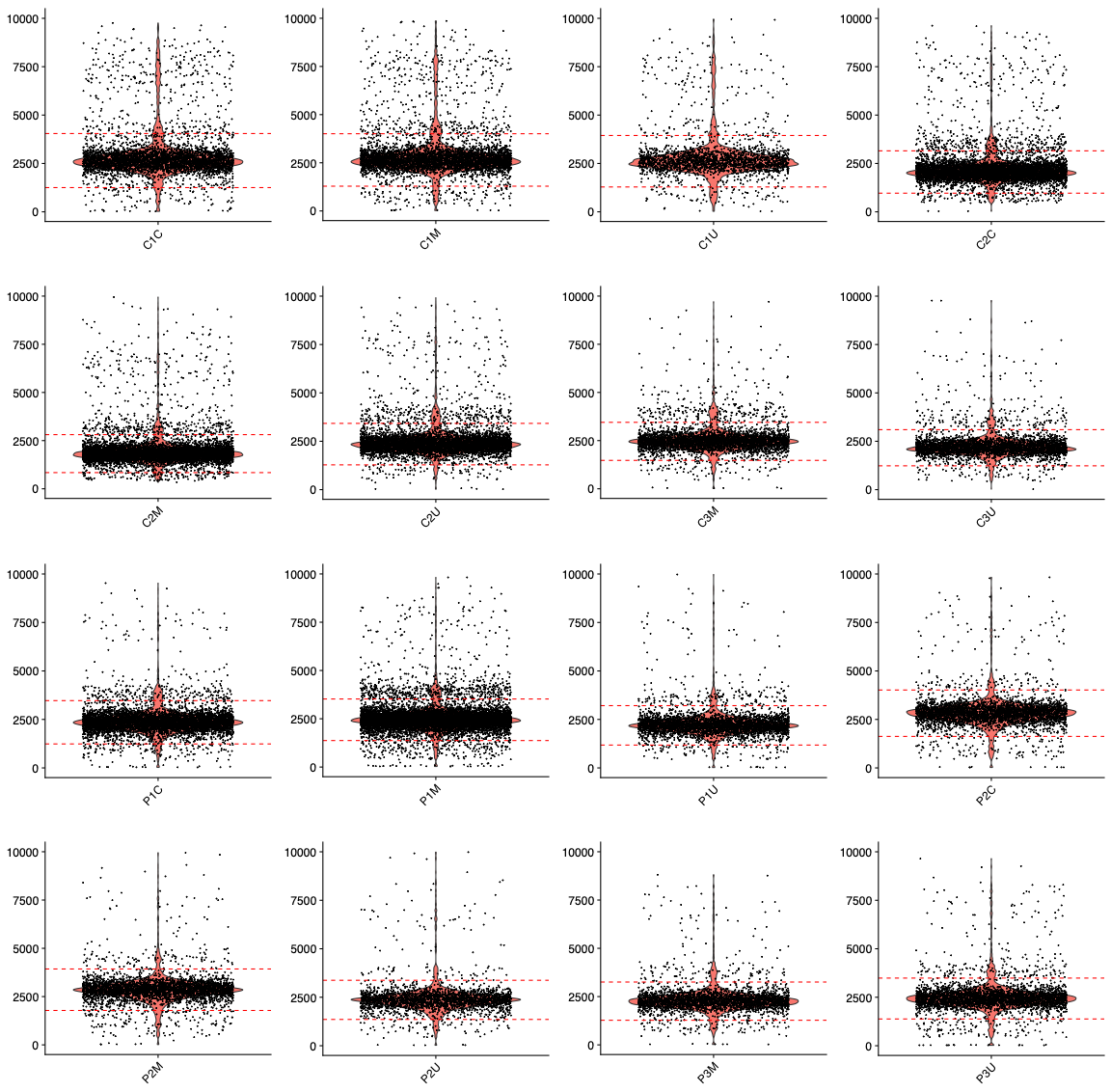

**Figure S2.** Distribution of the number of genes per cell for each individual sample. The red dotted line indicates the cutoff, defined as the median gene count per cell ± 3 median absolute deviations (MAD). Sample IDs follow the schema described in Figure S1.

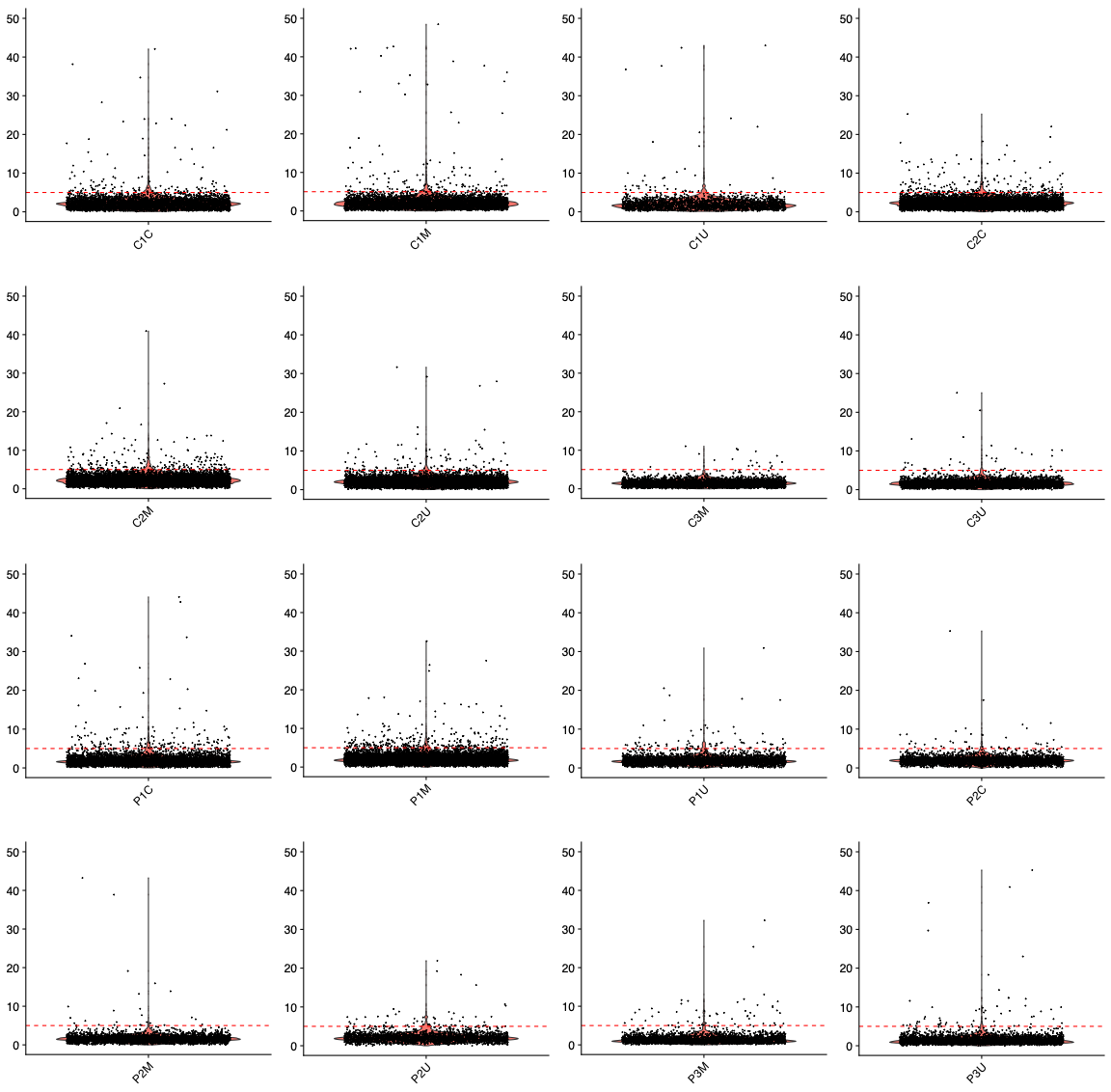

**Figure S3.** Distribution of the mitochondrial gene percentage (MT%) per cell for each individual sample. The red dotted line indicates the cutoff of MT% at 5%. Sample IDs follow the schema described in Figure S1.

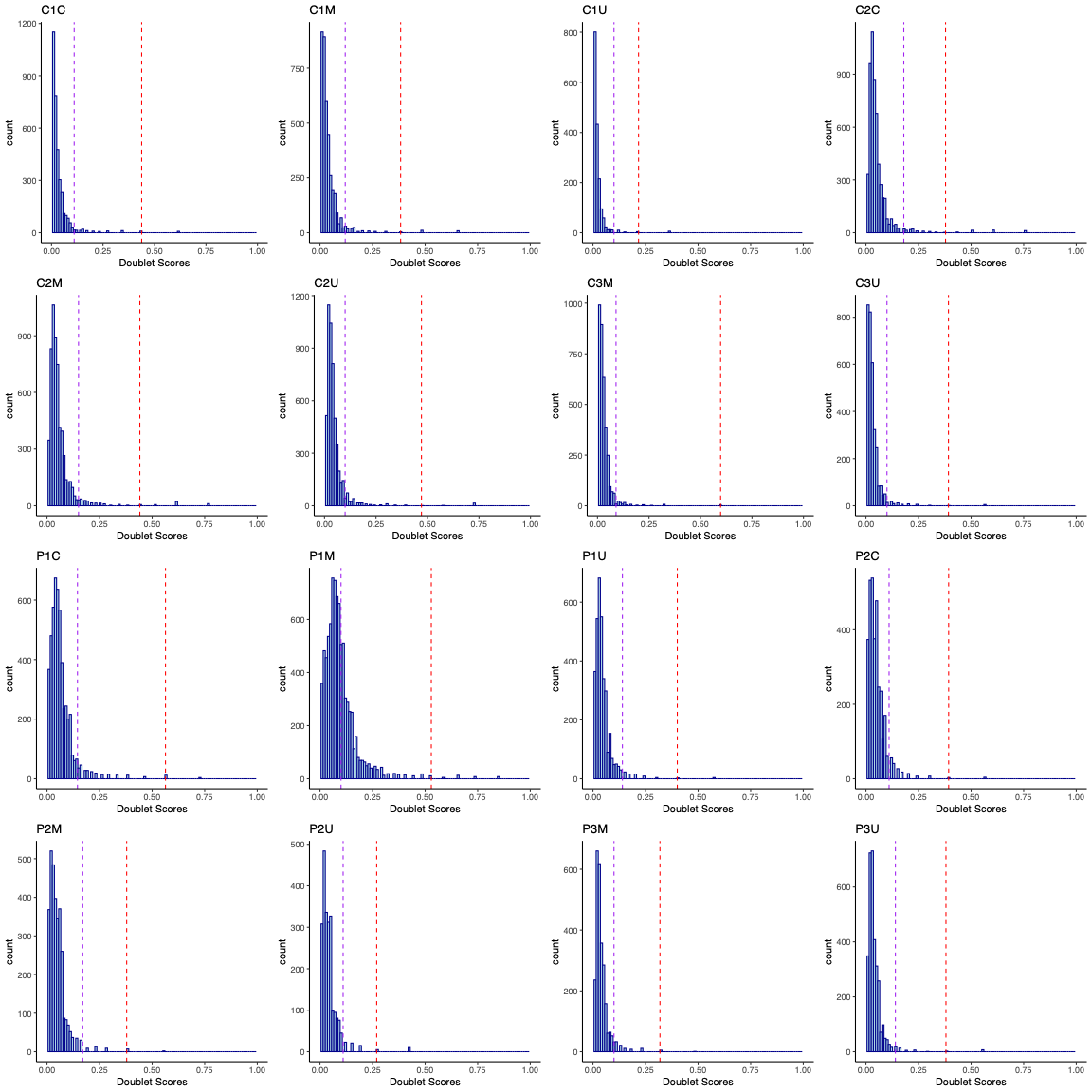

**Figure S4.** Distribution of the doublet score per cell for each individual sample estimated using “Scrublet”. Sample IDs follow the schema described in Figure S1. The red and purple dotted lines represent the automated (“Scrublet”) and manual conservative cutoffs, respectively, for selection of singlets.

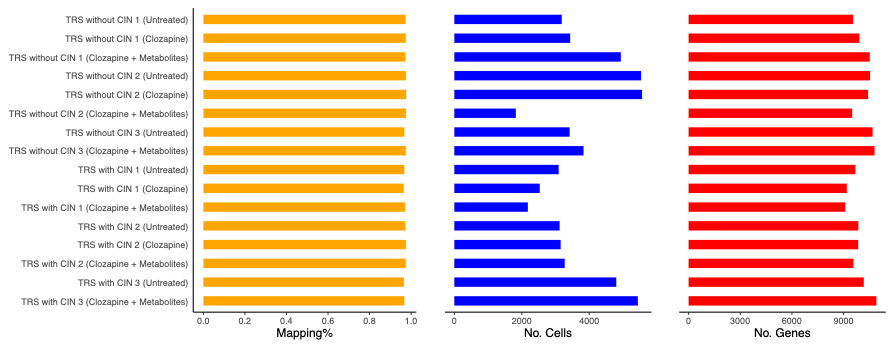

**Figure S5.** Distributions of mapping rates, cell count, and gene count per individual sample after quality control.

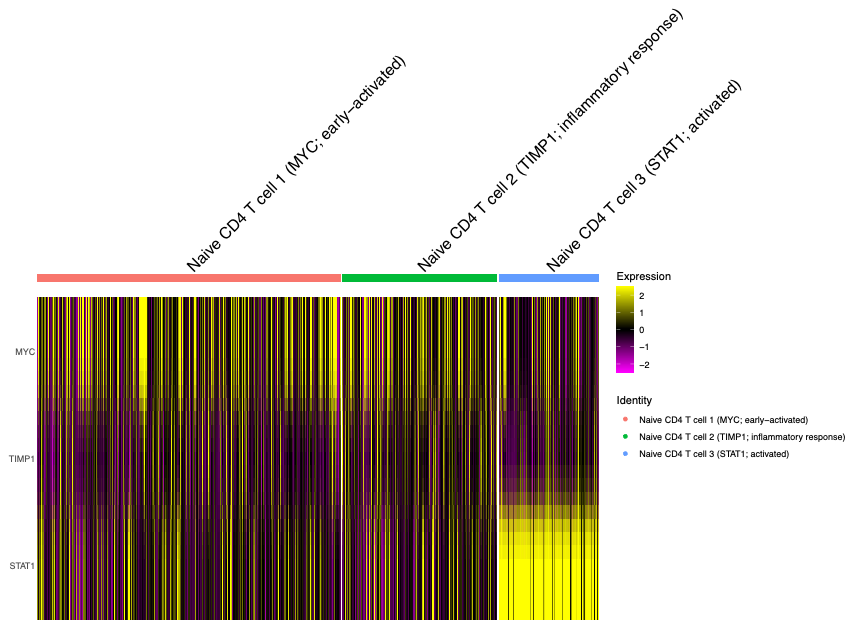

**Figure S6.** Normalised expression levels of the annotation marker gene for each naïve CD4 T cell subtype.

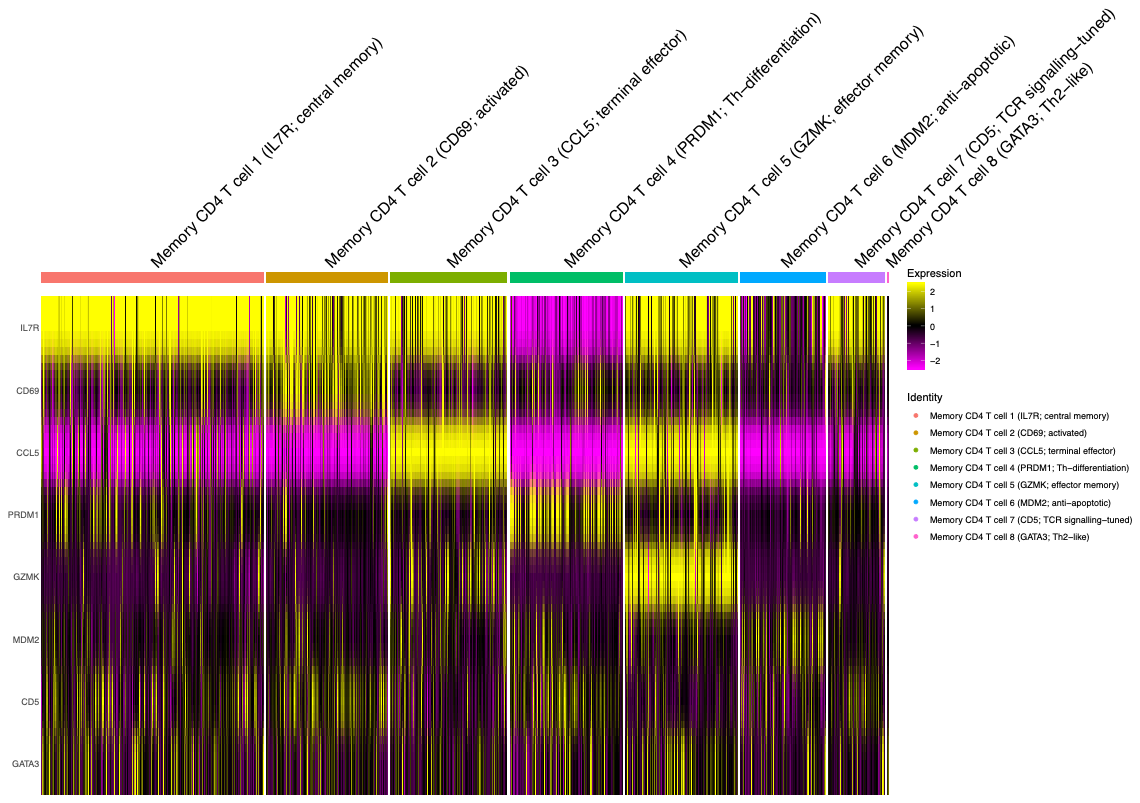

**Figure S7.** Normalised expression levels of the annotation marker gene for each memory CD4 T cell subtype.

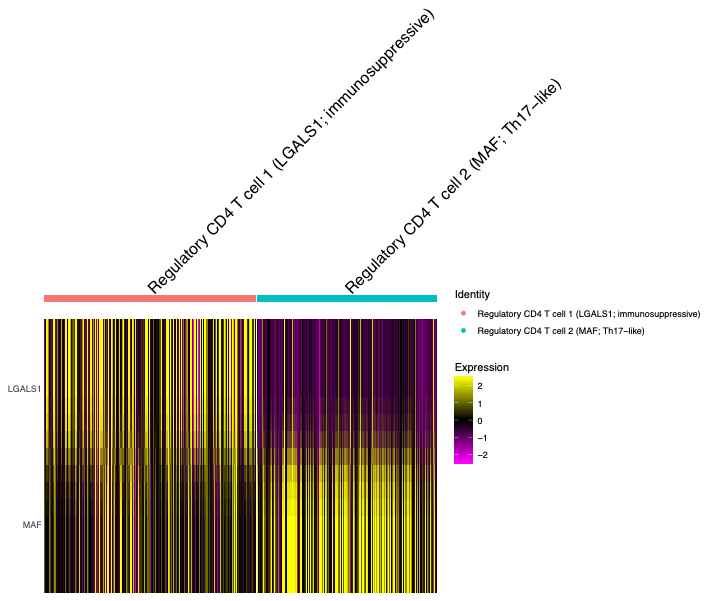

**Figure S8.** Normalised expression levels of the annotation marker gene for each regulatory CD4 T cell subtype.

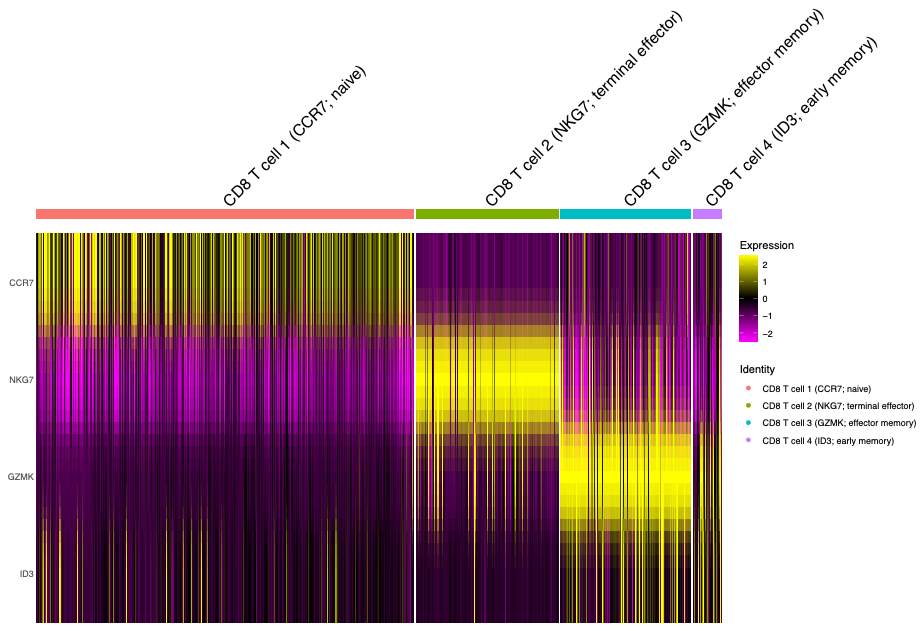

**Figure S9.** Normalised expression levels of the annotation marker gene for each CD8 T cell subtype.

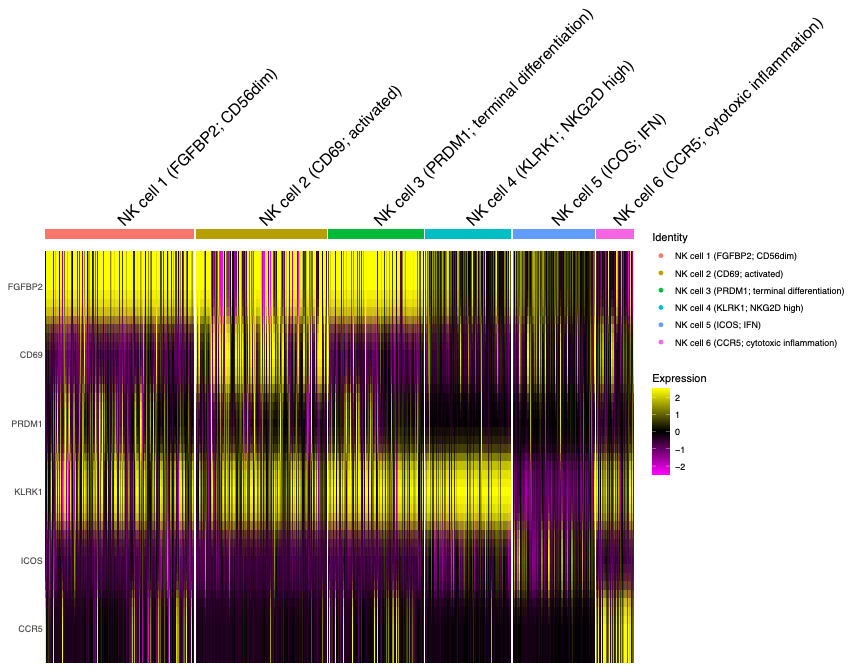

**Figure S10.** Normalised expression levels of the annotation marker gene for each NK cell subtype.

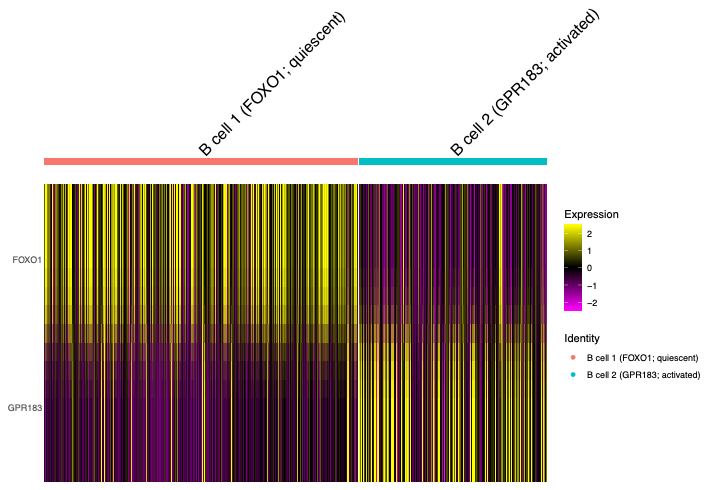

**Figure S11.** Normalised expression levels of the annotation marker gene for each B cell subtype.

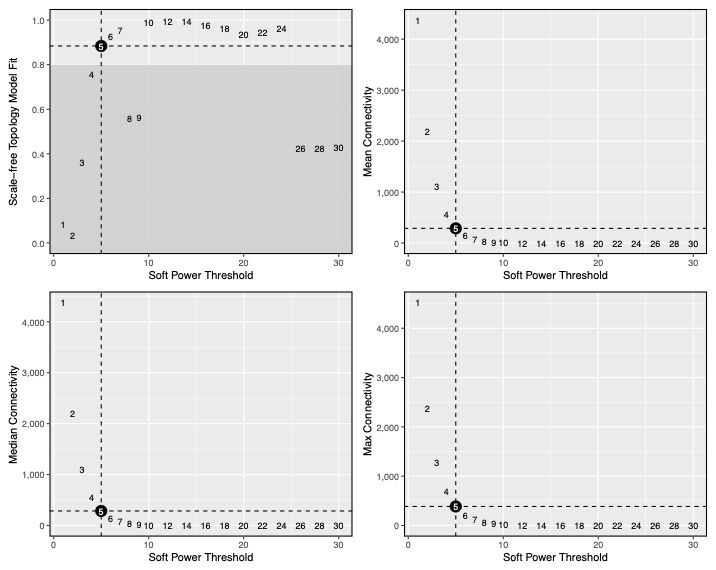

**Figure S12.** Soft‑threshold power selection for network construction in naive CD4 T cell 1 (early-activated). Relationships between the soft‑thresholding power and scale‑free topology model fit, mean connectivity, median connectivity, and maximum connectivity, respectively.

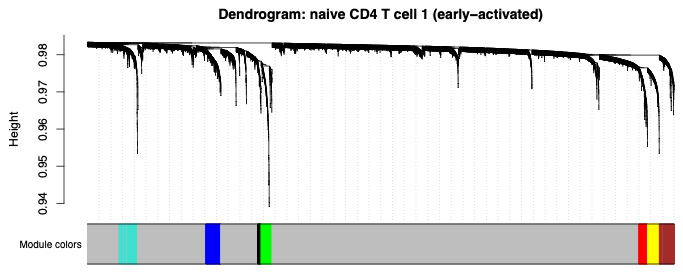

**Figure S13.** Gene co‑expression module detection in naive CD4 T cell 1 (early-activated). Hierarchical clustering dendrogram of genes constructed from the consensus topological overlap matrix. The module color bar beneath the dendrogram indicates gene modules identified by the dynamic tree‑cut procedure.

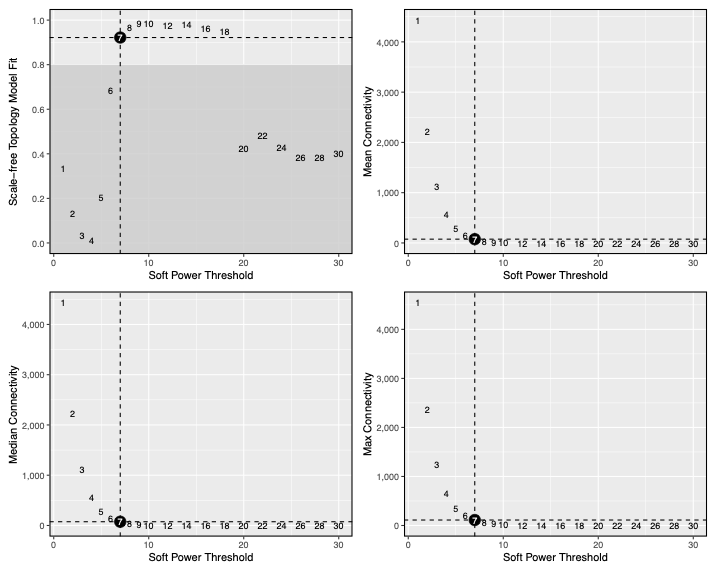

**Figure S14.** Soft‑threshold power selection for network construction in CD8 T cell 1 (naïve). Relationships between the soft‑thresholding power and scale‑free topology model fit, mean connectivity, median connectivity, and maximum connectivity, respectively.

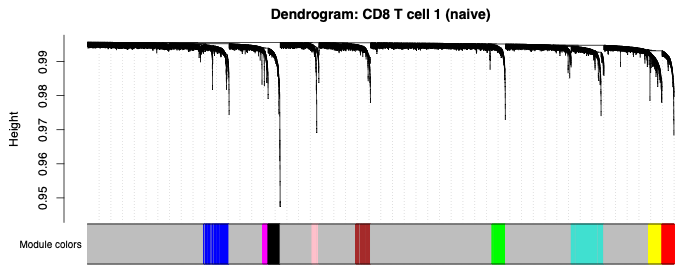

**Figure S15.** Gene co‑expression module detection in CD8 T cell 1 (naïve). Hierarchical clustering dendrogram of genes constructed from the consensus topological overlap matrix. The module color bar beneath the dendrogram indicates gene modules identified by the dynamic tree‑cut procedure.

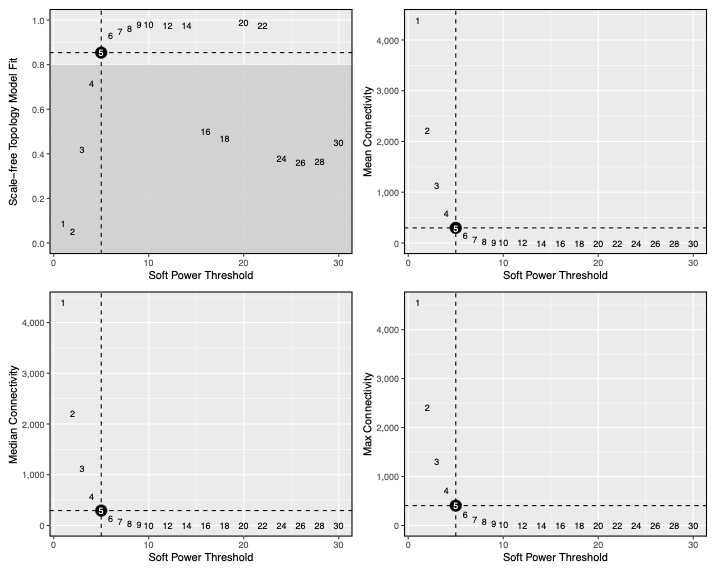

**Figure S16.** Soft‑threshold power selection for network construction in naive CD4 T cell 2 (inflammatory response). Relationships between the soft‑thresholding power and scale‑free topology model fit, mean connectivity, median connectivity, and maximum connectivity, respectively.

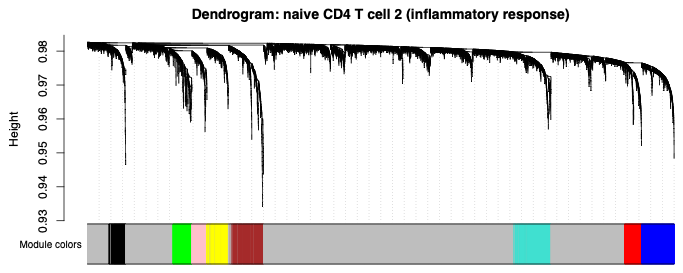

**Figure S17.** Gene co‑expression module detection in naive CD4 T cell 2 (inflammatory response). Hierarchical clustering dendrogram of genes constructed from the consensus topological overlap matrix. The module color bar beneath the dendrogram indicates gene modules identified by the dynamic tree‑cut procedure.

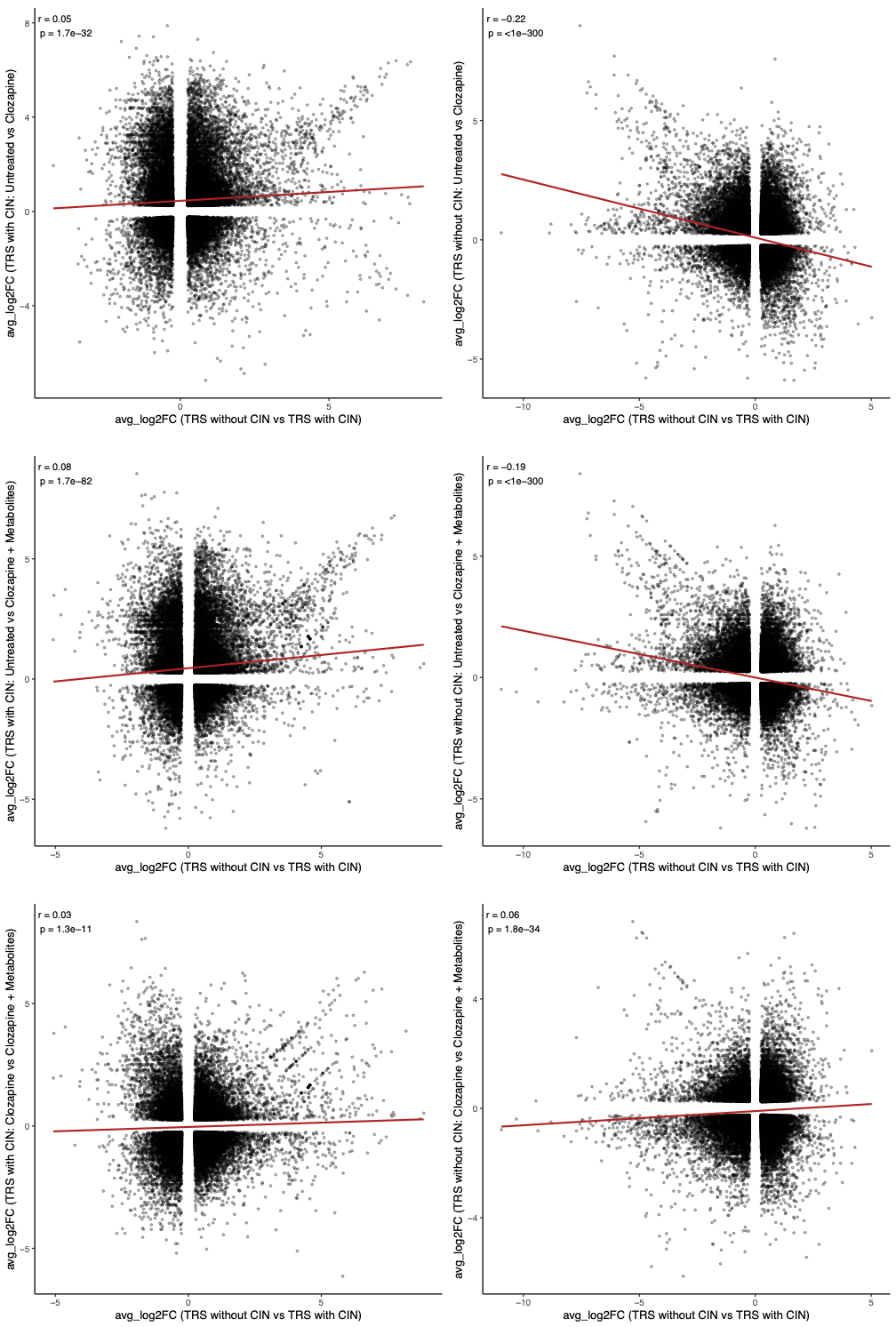

**Figure S18.** Scatter plots compare average log_2_FC per gene for the CIN gene set between TRS without CIN and TRS with CIN (x-axis) against treatment effects within TRS with CIN (left) or TRS without CIN (right) (y-axis) for three comparisons: “Untreated vs Clozapine”, “Untreated vs Clozapine + Metabolites”, and “Clozapine vs Clozapine + Metabolites”. Each point is one gene; Pearson correlations and p-values are shown in each panel.
